# Assessing multiplex tiling PCR sequencing approaches for detecting genomic variants of SARS-CoV-2 in municipal wastewater

**DOI:** 10.1101/2021.05.26.21257861

**Authors:** Xuan Lin, Melissa Glier, Kevin Kuchinski, Tenysha Ross-Van Mierlo, David McVea, John R. Tyson, Natalie Prystajecky, Ryan M. Ziels

## Abstract

Wastewater-based genomic surveillance of the SARS-CoV-2 virus shows promise to complement genomic epidemiology efforts. Multiplex tiled PCR is a desirable approach for targeted genome sequencing of SARS-CoV-2 in wastewater due to its low cost and rapid turnaround time. However, it is not clear how different multiplex tiled PCR primer schemes or wastewater sample matrices impact the resulting SARS-CoV-2 genome coverage. The objective of this work was to assess the performance of three different multiplex primer schemes, consisting of 150bp, 400bp, and 1200bp amplicons, as well as two wastewater sample matrices, influent wastewater and primary sludge, for targeted genome sequencing of SARS-CoV-2. Wastewater samples were collected weekly from five municipal wastewater treatment plants (WWTPs) in the Metro Vancouver region of British Columbia, Canada during a period of increased COVID-19 case counts from February to April, 2021. RNA extracted from clarified influent wastewater provided significantly higher genome coverage (breadth and median depth) than primary sludge samples across all primer schemes. Shorter amplicons appeared more resilient to sample RNA degradation, but were hindered by greater primer pool complexity in the 150bp scheme. The identified optimal primer scheme (400bp) and sample matrix (influent) was capable of detecting the emergence of mutations associated with genomic variants of concern, of which the daily wastewater load significantly correlated with clinical case counts. Taken together, these results provide guidance on best practices for implementing wastewater-based genomic surveillance, and demonstrate its ability to inform epidemiology efforts by detecting genomic variants of concern circulating within a geographic region.

**Importance:** Monitoring the genomic characteristics of the SARS-CoV-2 virus circulating in a population can shed important insights into epidemiological aspects of the COVID-19 outbreak. Sequencing every clinical patient sample in a highly populous area is a difficult feat, and thus sequencing SARS-CoV-2 RNA in municipal wastewater offers great promise to augment genomic surveillance by characterizing a pooled population sample matrix, particularly during an escalating outbreak. Here, we assess different approaches and sample matrices for rapid targeted genome sequencing of SARS-CoV-2 in municipal wastewater. We demonstrate that the optimal approach is capable of detecting the emergence of SARS-CoV-2 genomic variants of concern, with strong correlations to clinical case data in the province of British Columbia. These results provide guidance on best practices on, as well as further support for, the application of wastewater genomic surveillance as a tool to augment current genomic epidemiology efforts.

## Observation

Genomic surveillance of the SARS-CoV-2 virus plays a critical role in tracking its evolution during the current global COVID-19 pandemic (1–3). Recently, several emerging lineages of SARS-CoV-2, so-called variants of concern (VoCs), have been associated with increased levels of transmission (4), disease severity (5), and/or immune escape (6, 7). These VoCs have originated from various locations globally (4, 8), but are spreading within new geographic regions due to travel-associated and local transmission (9). Providing rapid detection of VoC infections within a population could thus help to inform effective public health outbreak mitigation strategies.

As the SARS-CoV-2 virus is shed in feces during infection (10), viral genome fragments can be detected in municipal wastewater, and have been associated with clinical case numbers within contributing regions (11–14). Previous work has demonstrated the potential to sequence SARS-CoV-2 fragments in municipal wastewater and detect single nucleotide variants (SNVs) that correspond to clinical cases in the contributing sewershed (15–17). As SARS-CoV-2 titers in wastewater are relatively low (11, 13), an enrichment step is typically needed prior to sequencing to improve sensitivity (15). The two main approaches for enriching SARS-CoV-2 RNA in wastewater include oligonucleotide based capture (15), and multiplex tiled PCR based targeted amplification (16, 17). The latter approach is promising for wastewater-based viral genomic surveillance due to its lower reagent cost and potential to be deployed rapidly and in remote locations (18). An important consideration for applying multiplex tiled PCR is the average amplicon length, as this can impact assay sensitivity in the case of RNA degradation (19). This could be particularly important for its application to wastewater based epidemiology, as SARS-CoV-2 particles and free RNA can undergo variable levels of degradation (20, 21), and may vary based on the type of wastewater sample matrix (e.g. influent versus primary sludge) (22). We therefore hypothesized that there may be an optimal tiled PCR amplicon size and wastewater sample matrix type that enables adequate genome coverage of SARS-CoV-2 for the identification of genomic VoCs.

### Wastewater sample matrix and multiplex tiled PCR amplicon length impact SARS-CoV-2 genome coverage

We sequenced a total of 96 wastewater samples collected between February 7^th^ to April 18^th^ 2021 across five municipal WWTPs in Vancouver, Canada using three different primer schemes for multiplex tiled PCR of SARS-CoV-2: Swift Bioscience’s 150bp amplicon scheme (n = 10 total, 3 sludge and 7 influent), ARTIC 400bp amplicon scheme (23) (n = 62 total, 8 sludge and 54 influent), and Freed/midnight 1200bp amplicon scheme (24) (n = 24 total, 4 sludge and 20 influent) (detailed methods in Text S1). Sludge samples failed to produce libraries with over 32% breadth of genome coverage across all primer schemes and sample cycle thresholds (C_t_’s) (Figure 1a-c). Conversely, influent wastewater samples produced libraries that had significantly higher breadth of coverage across all primer schemes (p<0.01, Tukey Test; Figure 1). One possible explanation for this finding could be that the sludge matrix was inhibitory to RT-PCR (11); however, no inhibition of RT-qPCR on sludge RNA extracts was detected using internal controls (Text S1, Table S2). Another potential reason for the lower genome coverage in sludge is that SARS-CoV-2 was more nonintact or its RNA more degraded with the direct sludge extraction compared to ultrafiltration of influent wastewater, as has been previously hypothesized (22). A third potential cause of discrepancies in genome coverage between sludge and influent wastewater samples could be higher off-target amplification in sludge extracts. Correspondingly, the sample type significantly impacted read mapping rates for all schemes after accounting for C_t_ values (p<0.01, two-way ANCOVA), with mean mapping rates of sludge samples being over 100-times lower than that of influent samples (0.01% vs. 11.3%, respectively; Table S1). Therefore, ultrafiltration of influent wastewater provided more suitable RNA extracts for multiplexed tiled PCR of SARS-CoV-2 than did direct extraction from wastewater sludge, likely due to a combination of greater SARS-CoV-2 RNA degradation and off-target amplification in sludge.

**Figure 1:**
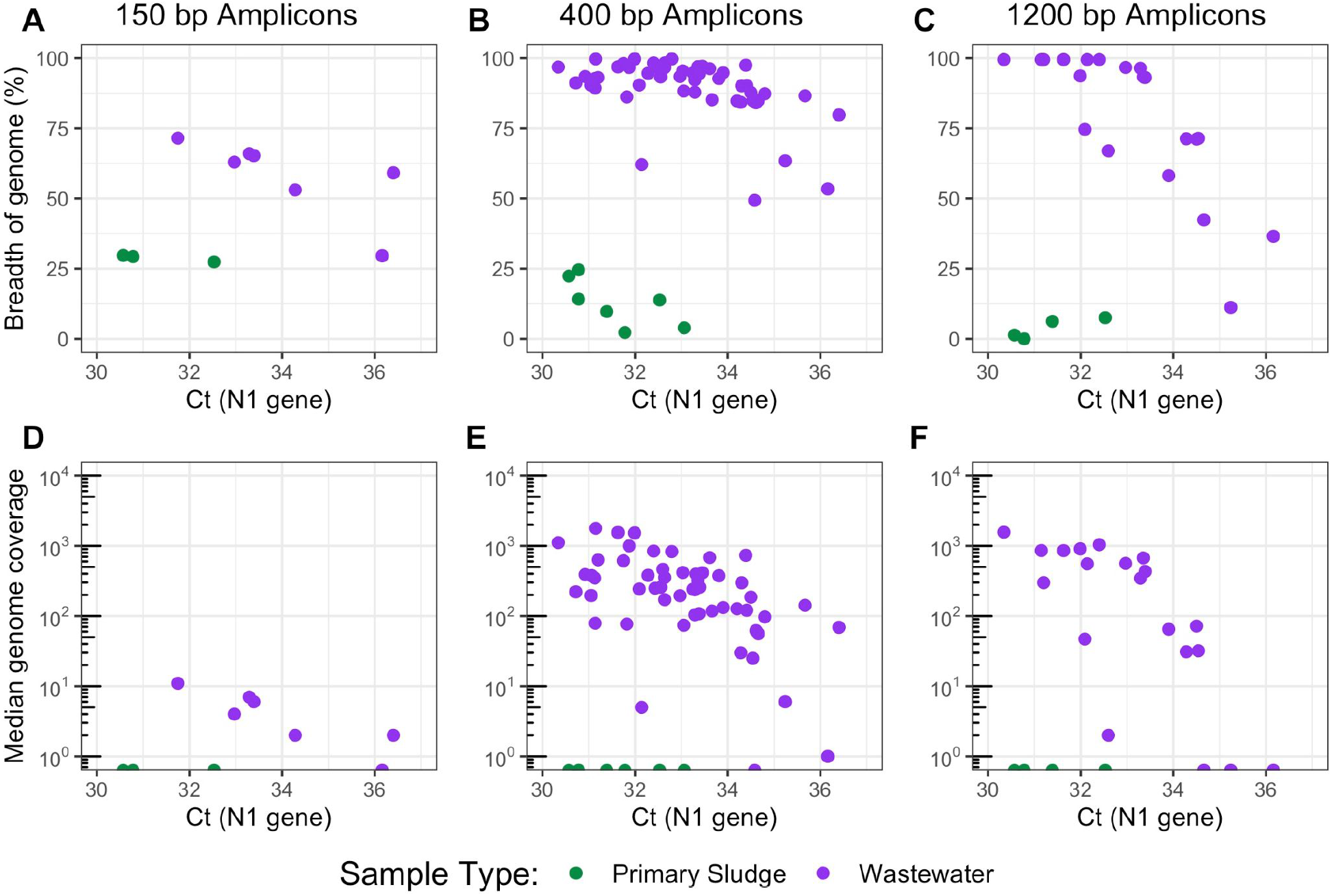
SARS-CoV-2 whole genome sequencing coverage results from three multiplex tiled PCR primer schemes, including breadth of genome coverage (over a read depth of 50) for (A) 150bp amplicons, (B) 400bp amplicons, and (C) 1200bp amplicons, as well as the median depth of coverage across the genome for (D) 150bp amplicons, (E) 400bp amplicons, and (F) 1200bp amplicons. Values are plotted versus the sample C_t_ value for the US CDC N1 assay, measured by RT-qPCR (see Text S1). Data points aligned with the x-axis (plots D-F) had values of zero, and could not be log-transformed.

If the level of RNA degradation within a wastewater sample impacts the resulting SARS-CoV-2 genome coverage, we would expect to see less of a drop-off in coverage at high C_t_’s for schemes with shorter amplicons. Indeed, we detected a significant effect of amplicon length on the breadth of genome coverage as a function of C_t_ (p<0.01, two-way ANCOVA). The median genome coverage with the 150bp amplicon scheme spanned one order of magnitude within influent wastewater samples with C_t_ values ranging from 31 to 37 (Figure 1d), while that from the 400bp and 1200bp schemes spanned 3.2 and 3.0 orders of magnitude, respectively (Figure 1e and 1f). Improvements with the 400bp versus the 1200bp scheme were marginal, yet 83% of paired influent samples with C_t_ values over 32.5 (10 of 12) showed higher breadth of coverage with the 400bp scheme (Figure S1). Thus, shorter amplicon schemes may be more robust to sample RNA degradation at higher C_t_ values. However, there was a tradeoff between amplicon length and genome coverage, as the magnitudes of the median genome coverage and breadth of coverage obtained with the 150bp scheme and influent samples were significantly lower than that of the 400bp scheme (p=0.022, 5.0e-9, respectively, Tukey Test). The lower breadth of coverage with the 150bp scheme could have been caused by more primer-primer interactions with a larger number of primers (19). Therefore, the 400bp primer scheme appears to strike a balance between resilience to sample RNA degradation and mitigating issues around primer pool complexity and multiplex amplicon balancing.

### SARS-CoV-2 whole genome sequencing from wastewater captures emergence of genomic variants in a geographic region

The sequence data produced via the 400bp primer scheme and influent wastewater samples was used to measure the frequency of VoC-associated SNVs (Table S3) across the five WWTPs over the study period. SNVs associated with the VoC lineages, B.1.1.7 and P.1, both increased to a maximum mean frequency of 60% across all WWTPs, respectively (Figure 2a, Figures S2-S4), while that of B.1.351 did not substantially increase (Figure S5). These findings align with the results clinical screening and sequencing of patient samples over the same period within the province of British Columbia, during which P.1 and B.1.1.7 became the dominant lineages while B.1.351 did not appreciably spread (25) (Figure 2b, Figures S3 and S5). At the time of publishing, VoC frequency data for clinical cases was only available at the provincial level; yet the health service areas corresponding to the 5 WWTP sewersheds accounted for 74% of total cases in the province during the study period (25). The flow-normalized daily loads of P.1 and B.1.1.7 across all WWTPs (in genome copies/day) were strongly correlated with clinical case counts of those lineages within the province for the corresponding epidemiological weeks (log_10_-log_10_ transformed, R^2^ = 0.89 and 0.87, respectively; Fig. 2c and Figure S3). The frequency of VoC-associated SNVs within influent wastewater measured with multiplex tiled PCR is therefore suitable to monitor community transmission of genomic variants within a sewershed. The onset of P.1- and B.1.1.7-associated SNVs within influent wastewater followed different patterns for the five WWTPs, providing additional support that wastewater SARS-CoV-2 sequencing can illuminate localized spread of genomic variants on a regional scale (15, 17). The rapid turnaround time (∼3 days from sampling to data generation here), low capital cost and high portability of nanopore sequencing combined with highly multiplexed tiled PCR for SARS-CoV-2 sequencing of wastewater shows great promise to complement genomic epidemiology efforts during the COVID-19 pandemic by detecting the emergence of VoCs within a pooled population sample.

**Figure 2:**
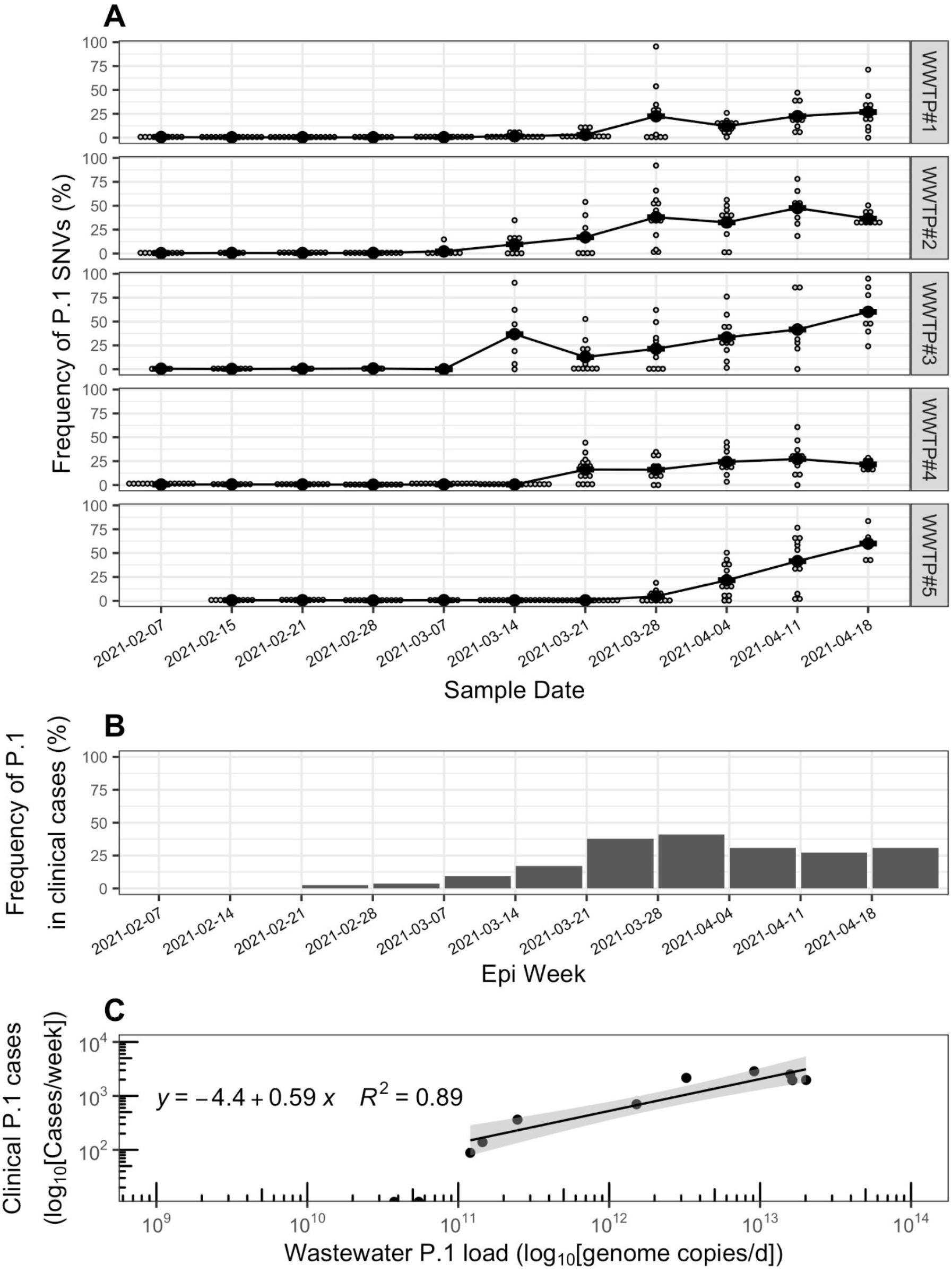
**(A)** Frequency of single nucleotide variants (SNVs) associated with the P.1 lineage of SARS-CoV-2 within influent wastewater samples from five wastewater treatment plants in Vancouver, British Columbia (BC), from February 7th to April 18th, 2021. Smaller grey dots represent the frequency of SNVs on sample dates, while the larger black points represent the mean across all detected SNVs. Only genome positions with a read coverage over 50 are included in SNV frequency calculations. **(B)** Frequency of the P.1 lineage in clinical COVID-19 patient cases in the province of BC, Canada over the study period. The frequencies in clinical patient cases correspond to an average value detected over an epidemiology (epi) week, and were adapted from (25). **(C)** Correlation between the wastewater cumulative daily load of P.1 genomes summed across all five WWTPs and the total P.1 clinical cases in the province of BC observed within the same epidemiological week. The wastewater P.1 daily load (genome copies/d) was approximated by normalizing copies of the US CDC N1 gene (copies/L) by daily flow rates (L/d) to obtain N1 loads (copies/d) for all WWTPs, and multiplying that by the mean frequency of P.1-associated SNVs in each WWTP across all sample dates. For each date, the cumulative P.1 daily load was determined by summing the P.1 loads across all five WWTPs. The P.1 clinical case counts by week were adapted from (25) by multiplying total provincial COVID-19 case counts by the frequency of P.1 in clinical provincial cases. Data points with zero clinical cases are shown aligned to the x-axis.

## Data Availability

The raw reads associated with all samples are available in the Short Read Archive under BioProject PRJNA731975. The accession numbers for each sample are also provided in Table S1, along with the sample metadata.

## Acknowledgements

We would like to thank Farida Bishay, Rob MacArthur, Daisy Espinosa and Alvin Louie, and the entire Metro Vancouver Environmental Management & Quality Control WWTP Laboratory Staff for collecting and delivering wastewater samples for this study and providing sample metadata. We would also like to thank Ziwen Ran for help with method development, and Matthias Krushel for help with processing wastewater samples. We also thank the Molecular and Microbial Genomics and Environmental Microbiology Laboratories at BCCDC Public Health Laboratory for materials and access to testing equipment, and the BCCDC and BC Regional Health Authorities for publicly sharing data on clinical case counts and variants of concern. This work was funded by the Natural Sciences and Engineering Research Council of Canada (NSERC, Alliance Grant, ALLRP 554612-20), BCCDC Foundation, Metro Vancouver, and Innovate BC.

## Notes

### Competing Interest Statement

The authors have declared no competing interest.

### Author Declarations

It was determined that no IRB or oversight body was required for this study.

## References

1. Rambaut A, Holmes EC, O’Toole Á, Hill V, McCrone JT, Ruis C, du Plessis L, Pybus OG. 2020. A dynamic nomenclature proposal for SARS-CoV-2 lineages to assist genomic epidemiology. 11. Nature Microbiology 5:1403–1407.

2. Bedford T, Greninger AL, Roychoudhury P, Starita LM, Famulare M, Huang M-L, Nalla A, Pepper G, Reinhardt A, Xie H, Shrestha L, Nguyen TN, Adler A, Brandstetter E, Cho S, Giroux D, Han PD, Fay K, Frazar CD, Ilcisin M, Lacombe K, Lee J, Kiavand A, Richardson M, Sibley TR, Truong M, Wolf CR, Nickerson DA, Rieder MJ, Englund JA, Investigators‡ TSFS, Hadfield J, Hodcroft EB, Huddleston J, Moncla LH, Müller NF, Neher RA, Deng X, Gu W, Federman S, Chiu C, Duchin JS, Gautom R, Melly G, Hiatt B, Dykema P, Lindquist S, Queen K, Tao Y, Uehara A, Tong S, MacCannell D, Armstrong GL, Baird GS, Chu HY, Shendure J, Jerome KR. 2020. Cryptic transmission of SARS-CoV-2 in Washington state. Science 370:571–575.

3. Candido DS, Claro IM, Jesus JG de, Souza WM, Moreira FRR, Dellicour S, Mellan TA, Plessis L du, Pereira RHM, Sales FCS, Manuli ER, Thézé J, Almeida L, Menezes MT, Voloch CM, Fumagalli MJ, Coletti TM, Silva CAM da, Ramundo MS, Amorim MR, Hoeltgebaum HH, Mishra S, Gill MS, Carvalho LM, Buss LF, Prete CA, Ashworth J, Nakaya HI, Peixoto PS, Brady OJ, Nicholls SM, Tanuri A, Rossi ÁD, Braga CKV, Gerber AL, Guimarães AP de C, Gaburo N, Alencar CS, Ferreira ACS, Lima CX, Levi JE, Granato C, Ferreira GM, Francisco RS, Granja F, Garcia MT, Moretti ML, Perroud MW, Castiñeiras TMPP, Lazari CS, Hill SC, Santos AA de S, Simeoni CL, Forato J, Sposito AC, Schreiber AZ, Santos MNN, Sá CZ de, Souza RP, Resende-Moreira LC, Teixeira MM, Hubner J, Leme PAF, Moreira RG, Nogueira ML, Brazil-UK Centre for Arbovirus Discovery D, Ferguson NM, Costa SF, Proenca-Modena JL, Vasconcelos ATR, Bhatt S, Lemey P, Wu C-H, Rambaut A, Loman NJ, Aguiar RS, Pybus OG, Sabino EC, Faria NR. 2020. Evolution and epidemic spread of SARS-CoV-2 in Brazil. Science 369:1255–1260.

4. Davies NG, Abbott S, Barnard RC, Jarvis CI, Kucharski AJ, Munday JD, Pearson CAB, Russell TW, Tully DC, Washburne AD, Wenseleers T, Gimma A, Waites W, Wong KLM, Zandvoort K van, Silverman JD, Group1‡ CC-19 W, Consortium‡ C-19 GU (COG-U, Diaz-Ordaz K, Keogh R, Eggo RM, Funk S, Jit M, Atkins KE, Edmunds WJ. 2021. Estimated transmissibility and impact of SARS-CoV-2 lineage B.1.1.7 in England. Science 372.

5. Davies NG, Jarvis CI, Edmunds WJ, Jewell NP, Diaz-Ordaz K, Keogh RH. 2021. Increased mortality in community-tested cases of SARS-CoV-2 lineage B.1.1.7. Nature 1–5.

6. Wibmer CK, Ayres F, Hermanus T, Madzivhandila M, Kgagudi P, Oosthuysen B, Lambson BE, de Oliveira T, Vermeulen M, van der Berg K, Rossouw T, Boswell M, Ueckermann V, Meiring S, von Gottberg A, Cohen C, Morris L, Bhiman JN, Moore PL. 2021. SARS-CoV-2 501Y.V2 escapes neutralization by South African COVID-19 donor plasma. 4. Nature Medicine 27:622–625.

7. Wang P, Nair MS, Liu L, Iketani S, Luo Y, Guo Y, Wang M, Yu J, Zhang B, Kwong PD, Graham BS, Mascola JR, Chang JY, Yin MT, Sobieszczyk M, Kyratsous CA, Shapiro L, Sheng Z, Huang Y, Ho DD. 2021. Antibody resistance of SARS-CoV-2 variants B.1.351 and B.1.1.7. Nature 1–6.

8. Tegally H, Wilkinson E, Giovanetti M, Iranzadeh A, Fonseca V, Giandhari J, Doolabh D, Pillay S, San EJ, Msomi N, Mlisana K, Gottberg A von, Walaza S, Allam M, Ismail A, Mohale T, Glass AJ, Engelbrecht S, Zyl GV, Preiser W, Petruccione F, Sigal A, Hardie D, Marais G, Hsiao M, Korsman S, Davies M-A, Tyers L, Mudau I, York D, Maslo C, Goedhals D, Abrahams S, Laguda-Akingba O, Alisoltani-Dehkordi A, Godzik A, Wibmer CK, Sewell BT, Lourenço J, Alcantara LCJ, Pond SLK, Weaver S, Martin D, Lessells RJ, Bhiman JN, Williamson C, Oliveira T de. 2020. Emergence and rapid spread of a new severe acute respiratory syndrome-related coronavirus 2 (SARS-CoV-2) lineage with multiple spike mutations in South Africa. medRxiv 2020.12.21.20248640.

9. Washington NL, Gangavarapu K, Zeller M, Bolze A, Cirulli ET, Schiabor Barrett KM, Larsen BB, Anderson C, White S, Cassens T, Jacobs S, Levan G, Nguyen J, Ramirez JM, Rivera-Garcia C, Sandoval E, Wang X, Wong D, Spencer E, Robles-Sikisaka R, Kurzban E, Hughes LD, Deng X, Wang C, Servellita V, Valentine H, De Hoff P, Seaver P, Sathe S, Gietzen K, Sickler B, Antico J, Hoon K, Liu J, Harding A, Bakhtar O, Basler T, Austin B, MacCannell D, Isaksson M, Febbo PG, Becker D, Laurent M, McDonald E, Yeo GW, Knight R, Laurent LC, de Feo E, Worobey M, Chiu CY, Suchard MA, Lu JT, Lee W, Andersen KG. 2021. Emergence and rapid transmission of SARS-CoV-2 B.1.1.7 in the United States. Cell https://doi.org/10.1016/j.cell.2021.03.052.

10. Pan Y, Zhang D, Yang P, Poon LLM, Wang Q. 2020. Viral load of SARS-CoV-2 in clinical samples. The Lancet Infectious Diseases 20:411–412.

11. D’Aoust PM, Mercier E, Montpetit D, Jia J-J, Alexandrov I, Neault N, Baig AT, Mayne J, Zhang X, Alain T, Langlois M-A, Servos MR, MacKenzie M, Figeys D, MacKenzie AE, Graber TE, Delatolla R. 2021. Quantitative analysis of SARS-CoV-2 RNA from wastewater solids in communities with low COVID-19 incidence and prevalence. Water Research 188:116560.

12. Wolfe MK, Archana A, Catoe D, Coffman MM, Dorevich S, Graham KE, Kim S, Grijalva LM, Roldan-Hernandez L, Silverman AI, Sinnott-Armstrong N, Vugia DJ, Yu AT, Zambrana W, Wigginton KR, Boehm AB. 2021. Scaling of SARS-CoV-2 RNA in Settled Solids from Multiple Wastewater Treatment Plants to Compare Incidence Rates of Laboratory-Confirmed COVID-19 in Their Sewersheds. Environ Sci Technol Lett https://doi.org/10.1021/acs.estlett.1c00184.

13. Peccia J, Zulli A, Brackney DE, Grubaugh ND, Kaplan EH, Casanovas-Massana A, Ko AI, Malik AA, Wang D, Wang M, Warren JL, Weinberger DM, Arnold W, Omer SB. 2020. Measurement of SARS-CoV-2 RNA in wastewater tracks community infection dynamics. 10. Nature Biotechnology 38:1164–1167.

14. Wu F, Zhang J, Xiao A, Gu X, Lee WL, Armas F, Kauffman K, Hanage W, Matus M, Ghaeli N, Endo N, Duvallet C, Poyet M, Moniz K, Washburne AD, Erickson TB, Chai PR, Thompson J, Alm EJ. 2020. SARS-CoV-2 Titers in Wastewater Are Higher than Expected from Clinically Confirmed Cases. mSystems 5.

15. Crits-Christoph A, Kantor RS, Olm MR, Whitney ON, Al-Shayeb B, Lou YC, Flamholz A, Kennedy LC, Greenwald H, Hinkle A, Hetzel J, Spitzer S, Koble J, Tan A, Hyde F, Schroth G, Kuersten S, Banfield JF, Nelson KL. 2021. Genome Sequencing of Sewage Detects Regionally Prevalent SARS-CoV-2 Variants. mBio 12.

16. Nemudryi A, Nemudraia A, Wiegand T, Surya K, Buyukyoruk M, Cicha C, Vanderwood KK, Wilkinson R, Wiedenheft B. 2020. Temporal Detection and Phylogenetic Assessment of SARS-CoV-2 in Municipal Wastewater. Cell Reports Medicine 1:100098.

17. Izquierdo-Lara R, Elsinga G, Heijnen L, Munnink BBO, Schapendonk CME, Nieuwenhuijse D, Kon M, Lu L, Aarestrup FM, Lycett S, Medema G, Koopmans MPG, Graaf M de. Monitoring SARS-CoV-2 Circulation and Diversity through Community Wastewater Sequencing, the Netherlands and Belgium - Volume 27, Number 5—May 2021 - Emerging Infectious Diseases journal - CDC https://doi.org/10.3201/eid2705.204410.

18. Quick J, Loman NJ, Duraffour S, Simpson JT, Severi E, Cowley L, Bore JA, Koundouno R, Dudas G, Mikhail A, Ouédraogo N, Afrough B, Bah A, Baum JHJ, Becker-Ziaja B, Boettcher JP, Cabeza-Cabrerizo M,Camino-Sánchez Á, Carter LL, Doerrbecker J, Enkirch T, Dorival IG-, Hetzelt N, Hinzmann J, Holm T, Kafetzopoulou LE, Koropogui M, Kosgey A, Kuisma E, Logue CH, Mazzarelli A, Meisel S, Mertens M, Michel J, Ngabo D, Nitzsche K, Pallasch E, Patrono LV, Portmann J, Repits JG, Rickett NY, Sachse A, Singethan K, Vitoriano I, Yemanaberhan RL, Zekeng EG, Racine T, Bello A, Sall AA, Faye O, Faye O, Magassouba N, Williams CV, Amburgey V, Winona L, Davis E, Gerlach J, Washington F, Monteil V, Jourdain M, Bererd M, Camara A, Somlare H, Camara A, Gerard M, Bado G, Baillet B, Delaune D, Nebie KY, Diarra A, Savane Y, Pallawo RB, Gutierrez GJ, Milhano N, Roger I, Williams CJ, Yattara F, Lewandowski K, Taylor J, Rachwal P, J. Turner D, Pollakis G, Hiscox JA, Matthews DA, Shea MKO, Johnston AM, Wilson D, Hutley E, Smit E, Di Caro A, Wölfel R, Stoecker K, Fleischmann E, Gabriel M, Weller SA, Koivogui L, Diallo B, KeÏta S, Rambaut A, Formenty P, Günther S, Carroll MW. 2016. Real-time, portable genome sequencing for Ebola surveillance. 7589. Nature 530:228–232.

19. Quick J, Grubaugh ND, Pullan ST, Claro IM, Smith AD, Gangavarapu K, Oliveira G, Robles-Sikisaka R, Rogers TF, Beutler NA, Burton DR, Lewis-Ximenez LL, de Jesus JG, Giovanetti M, Hill SC, Black A, Bedford T, Carroll MW, Nunes M, Jr LCA, Sabino EC, Baylis SA, Faria NR, Loose M, Simpson JT, Pybus OG, Andersen KG, Loman NJ. 2017. Multiplex PCR method for MinION and Illumina sequencing of Zika and other virus genomes directly from clinical samples 16.

20. Bivins A, Greaves J, Fischer R, Yinda KC, Ahmed W, Kitajima M, Munster VJ, Bibby K. 2020. Persistence of SARS-CoV-2 in Water and Wastewater. Environ Sci Technol Lett 7:937–942.

21. Wurtzer S, Waldman P, Ferrier-Rembert A, Frenois-Veyrat G, Mouchel JM, Boni M, Maday Y, Marechal V, Moulin L. 2021. Several forms of SARS-CoV-2 RNA can be detected in wastewaters: implication for wastewater-based epidemiology and risk assessment. Water Research 117183.

22. Kantor RS, Nelson KL, Greenwald HD, Kennedy LC. 2021. Challenges in Measuring the Recovery of SARS-CoV-2 from Wastewater. Environ Sci Technol 55:3514–3519.

23. Tyson JR, James P, Stoddart D, Sparks N, Wickenhagen A, Hall G, Choi JH, Lapointe H, Kamelian K, Smith AD, Prystajecky N, Goodfellow I, Wilson SJ, Harrigan R, Snutch TP, Loman NJ, Quick J. 2020. Improvements to the ARTIC multiplex PCR method for SARS-CoV-2 genome sequencing using nanopore. bioRxiv https://doi.org/10.1101/2020.09.04.283077.

24. Freed NE, Vlková M, Faisal MB, Silander OK. 2020. Rapid and inexpensive whole-genome sequencing of SARS-CoV-2 using 1200 bp tiled amplicons and Oxford Nanopore Rapid Barcoding. Biology Methods and Protocols 5.

25. BC Center for Disease Control. 2021. Weekly update on Variants of Concern (VOC). May 6, 2021

