## Supplemental Information for "Assessing multiplex tiling PCR sequencing approaches for detecting genomic variants of SARS-CoV-2 in municipal wastewater"

**Text S1: Supplemental Methods**

***Sample collection, concentration and RNA extraction***

Composite samples (24-hr flow-weighted) of raw influent wastewater were collected weekly from five WWTPs in Metro Vancouver, British Columbia, using an autosampler device (BVS4300C, Campbell Scientific, Edmonton, AB, Canada at WWTPs #1-4; 5800 Sampler, Teledyne ISCO, Lincoln, NE, USA at WWTP #5). Following collection, 1 L samples were aliquoted from the composite samples and directly shipped at 4°C and kept at 4°C for no more than 48 h before processing, in triplicate, to obtain viral concentrates. Solids and larger particles were removed by centrifugation of 15 mL aliquots in conical tubes at 4200 *x g* for 20 minutes at 4°C. The supernatant volume was measured, and then processed by centrifugal ultrafiltration using Amicon Ultra-15 Filters with a 10-kDa MWCO (Sigma-Aldrich Canada Co. Oakville, ON, Canada) at 4200 *x g* for 35 minutes at 4°C. This generated 170 to 250 µL of viral concentrate that was stored in DNA LoBind Tubes (Eppendorf Canada, Mississauga, ON, Canada) at -20°C for immediate use, or -80°C for later use. Nucleic acids were extracted instantaneously from ~200 µL of viral concentrate using the automated NucliSens easyMAG instrument with the NucliSens magnetic extraction reagents (bioMerieux Canada Inc., Saint-Laurent, QC, Canada) according to the instructions of the manufacturer. A volume of 100 µL of DNA/RNA eluate was selected and immediately used for detection, or was aliquoted into Eppendorf DNA LoBind Tubes at volumes ready-to-use for downstream sequencing and stored at -20°C for short-term storage or -80°C for long-term storage. A positive SARS-CoV-2 clinical specimen was used as the positive control and DEPC water (Invitrogen, Thermo Fisher Scientific, Waltham, MA, USA) as the negative control.

Composite samples of thickened primary sludge (TPS) samples (1.5 L) were collected, at all but one WWTP (WWTP #5 did not have a primary clarifier and so no samples were collected there), from the discharge of the primary gravity thickener, which processes the settled sludge from the primary sedimentation tank after grit removal and coarse screening. These composite primary sludge samples consisted of 500 mL grab samples collected three times daily from each of the primary gravity thickeners. Primary sludge samples were shipped at 4°C and kept at 4°C for no more than 24 h before extraction. The primary sludge was pelleted by centrifugation at 4°C and 10,000 x*g* for 30 minutes, and 250 mg of pelleted solids were then transferred to lysis tubes for RNA extraction using the Qiagen AllPrep PowerViral DNA/RNA Kit (Qiagen, Toronto, ON, Canada) following manufacturer's instructions.

***Reverse Transcription-quantitative PCR (RT-qPCR)***

SARS-CoV-2 RNA was detected using the N1 primer-probes published by the US CDC and manufactured by IDT (2019-nCoV RUO Kit, Integrated DNA Technologies, Coralville, IA, USA) and the Applied Biosystems TaqMan Fast Virus 1-Step Master Mix (Applied Biosystems, Fisher Scientific, Waltham, MA, USA). RT-qPCR was performed on an Applied Biosystems 7500 PCR instrument (Applied Biosystems, Fisher Scientific, Waltham, MA, USA). Thermal cycling was performed at 50°C for 5 minutes for reverse transcription, followed by 95°C for 20 seconds for enzyme activation and then 45 amplification cycles of 95°C for 3 seconds, 60°C for 30 seconds. Samples were run in technical triplicates in MicroAmp Fast Optical 96 well Reaction plates (0.1 mL) (Applied Biosystems, Fisher Scientific, Waltham, MA, USA) using 5 μL of template for a total reaction volume of 20 μL. The target threshold was manually set to 0.05. To obtain relative quantification, a six-point standard curve was prepared from 10-fold series of dilutions from 10 copies to $10^6$ copies of SARS-CoV-2 gBlock DNA gene fragments (Integrated DNA Technologies, Coralville, IA, USA) enclosing the RT-qPCR targets, and was run in triplicate. The standard calibration curve had a linear amplification range of 5 to 5 x $10^5$ copies per reaction, an efficiency of 96.93%, a slope of -3.3993, a y-intercept of 36.5893, and a coefficient of determination ($R^2$) of 0.99999. The DNA fragment sequences of the standards are provided in Supplemental Methods Table 1.

**Supplemental Methods Table 1 –** IDT Custom DNA fragment control oligo (gBlock) sequence used to create standard curves for relative quantification of SARS-CoV-2 RNA in wastewater samples.

| Name | Targets Contained | Sequence |
|---|---|---|
| SARS-CoV-2 All Targets | E_Sarbeco, US CDC N1 and N2, RdRp | TAT CTG GTG ATA CAT GAA CAG ATC CGT GCA CCG TCC<br>ATT CGT TTC GGA AGA AAC AGG TAC GTT AAT AGT TAA<br>TAG CGT ACT TCT TTT TCT TGC TTT CGT GGT ATT CTT<br>GCT AGT CAC ACT AGC CAT CCT TAC TGC GCT TCG ATT<br>GTG TGC GTA CTG CTG CAA TAT TGT TAA CGT GAG TTT<br>AGT AAC CCA AAG ACC ACA TTG GCA CCC GCA ATC CTA<br>ATA ACA ATG CTG CCA CCG TGC TAC AAC TTC CTC AAG<br>GAA CAA CAA AGT CAG CCT GCA TGT CTG ATA ATG GAC<br>CCC AAA ATC AGC GAA ATG CAC CCC GCA TTA CGT TTG<br>GTG GAC CCT CAG ATT CAA CTG GCA GTA ACC AGA ATG<br>GAG AAC GCA GTG GGG CGC GAT CAA AAC AAC GTC<br>GGC CCC AAG AGC TGT CAG CAC TAC TAA CTT GCG<br>GTC AGT ATG ATT CAA TGA GTT ATG AGG ATC AAG ATG<br>CAC TTT TCG CAT ATA CAA AAC GTA ATG TCA TCC CTA<br>CTA TAA CTC AAA TGA ATC TTA AGT ATG CCA TTA GTG<br>CAA AGA ATA GAG CTC GCA CCG TAG CTG GTG TCT CTA<br>TCT GTA GTA CTA TGA CCA ATA GAC AGT TTC ATC AAA<br>AAT TAT TGA AAT CAA TAG CCG CCA CTA GAG GAG CTA<br>CTG TAG TAA TTG GAA CAA GCA AAT TCT ATG GTG GTT<br>GGC ACA ACA TGT TAA AAA CTG TTT ATA GTG ATG TAG<br>AAA ACC CTC ACC TTA TGG GTT GGG ATT ATC CTA AAT<br>GTG ATA GAG CCA TGC CTA ACA TGC TTA GAA TTA TGG<br>CCT CAC TTG TTC TTG CTC GCA AAC ATA CAA CGT GTT<br>GTA GCT TGT CAC ACC GTT TCT ATA GAT TAG CTA ATG<br>AGT GTG CTC AAG TAT TGA GTG AAA TGG TCA TGT GTG<br>GCG GTT CAC TAT ATG TTA AAC CAG GTG GAA CCT CAT<br>CAG GAG ATG CCA CAA CTG CTT ATG CTA ATA GTG TTT<br>TTA ACA TTT GTC AAG CTG TCA CGG CCA ATG TTA ATG<br>CAC TTT TAT CTA CTG ATG GTA ACA AAA TTG CCG ATA<br>AGT ATG TCC GCA ATT TAC AAC ACA GAC TTT ATG AGT<br>GTC TCT ATA GAA ATA GAG ATG TTG ACA CAG ACT TTG<br>TGA ATG AGT TTT ACG CAT ATT TGC GTA AAC ATT TCT<br>CAA TGA TGA TAC TCT CTG ACG ATG CTG TTG TGT GTT<br>TCA ATA GCA CTT ATG CAT |

A positive SARS-CoV-2 clinical specimen was used as the positive control for both extraction and RT-qPCR. DEPC water (Invitrogen, Thermo Fisher Scientific, Waltham, MA, USA) served as the negative control for both extraction and RT-qPCR. Nuclease-free water (Invitrogen) was used as the no-template negative RT-qPCR control to assess contamination within the qPCR procedure. Influent wastewater samples were assessed for inhibition by spiking a known quantity of a west nile virus (WNV) Armoured RNA (Asuragen, Austin, TX, USA) into the lysis buffer for each sample during RNA extraction. Presence of inhibition was assessed by detecting WNV by RT-qPCR in the spiked samples and the spiked water control samples using the following primers and probe: NSF-2F (5'-GAAGAGACCTGCGGCTCATG), NSF-2R (5'-CGGTAGGGACCCAATTCACA), and probe NSF-2 Probe (5'-TYE 665- CCA ACG CCA TTT GCT CCG CTG -IBRQ). The WNV amplification scheme was the same as the US CDC N1 gene. Inhibition was defined as a delay of at least 3 cycles in the spiked sample as compared to a water control spiked with the same concentration of WNV Armoured RNA. We did not detect PCR inhibition with influent wastewater samples. Primary sludge samples were assessed for inhibition with: 1) Veto VetMax Internal Positive Control Assay (Applied Biosystems, Waltham, MA, USA) by adding the same concentration of the internal positive control (IPC) to all RT-qPCR reactions; and 2) addition of 5 mg/mL BSA (Invitrogen, Thermo Fisher Scientific, Waltham, MA, USA) to the RT-qPCR reactions. Presence of inhibition was defined as observing significantly higher $C_t$ values of the IPC in sludge samples compared to that in the positive control. No inhibition was detected using the VetMax IPC assay with primary sludge samples, nor was any impact of BSA observed on reducing sample $C_t$ values of the US CDC N1 assay (Table S2).

***Reverse Transcription and Multiplex Tiled PCR for Whole Genome Sequencing***

*Reverse Transcription and cDNA cleanup*

Extracted wastewater and primary sludge RNAs were reverse transcribed into complementary DNA (cDNA) using the SuperScript IV First-Strand Synthesis System (Invitrogen, Waltham, MA, USA). Reverse transcript reactions were comprised of 12 µL RNA input, 2.5 µM random hexamers, 0.5 mM dNTPs mix, 5 mM DTT, 2.0U/µL Ribonuclease Inhibitor, and 1x 5x SSIV Buffer in 40 µL reaction volume. Input RNAs were first incubated in a MiniAmp™ Plus Thermal Cycler (Applied Biosystems, Waltham, MA, USA) with random hexamers and dNTPs at 65°C for 5 min, then cooled on ice for 1 min before the addition of reverse transcript reaction mix. The combined reaction mixtures were incubated in a MiniAmp™ Plus Thermal Cycler (Applied Biosystems, Waltham, MA, USA) at 42 °C for 50 min, followed by enzyme inactivation at 70 °C for 10 min. The remaining RNAs were hydrolyzed by adding 8 µL 0.5M EDTA (pH=8.0) and 8 µL 1N NaOH to the reverse transcription reaction mixture and incubated in a MiniAmp™ Plus Thermal Cycler (Applied Biosystems, Waltham, MA, USA) at 65 °C for 15 min. This hydrolysis reaction was cleaned with Zymo DNA Clean & Concentrator-5 kit following the manufacturer's instructions (Zymo Research, Irvine, CA, USA).

Primer stocks for the 400 bp ARTIC V3 primer scheme (1) and the 1200 bp Freed/'midnight' primer scheme (2) panel were prepared by diluting the 100 µM pools (Integrated DNA Technologies, Coralville, IA, USA; File S1) 1:10 in nuclease-free water (Thermo Fisher Scientific, Waltham, MA, USA). Primer sequences of the 150 bp panel were obtained from the Swift Biosciences product description website (https://swiftbiosci.com/swift-amplicon-sars-cov-2-panel/) (Swift Biosciences, Ann Arbor, MI, USA). The primers were organized into two primer pools of non-overlapping tiled amplicons (File S1). The primers were produced by Integrated DNA Technologies, dissolved in IDTE (0.1 mM EDTA and 10 mM Tris) at a stock concentration of 60 nM per primer. EDTA was removed from the primers by cleaning with the Zymo Oligo Clean & Concentrator kit following the manufacturer's instructions (Zymo Research, Irvine, CA, USA), and resuspending in nuclease-free water to a stock concentration of 60 nM per primer.

*Multiplex Tiled PCR of SARS-CoV-2*

Cleaned cDNA was prepared for whole genome sequencing following amplification with three multiplex tiled PCR schemes (File S1): (i) 150 bp amplicons based on the Swift Amplicon SARS-

CoV-2 Panel primer scheme (Swift Biosciences, Ann Arbor, MI, USA); (ii) 400 bp amplicons with the ARTIC V3 primer scheme, and (iii) 1200 bp amplicons with the Freed/'midnight' primer scheme. The metadata of the samples prepared with the three multiplex primer schemes are provided in Table S1. A no-template negative control (nuclease-free water), along with a positive control of synthetic SARS-CoV-2 RNA of the Wuhan-1 reference genome (102019, Twist Control-1, GenBank ID: MT007544.1, Twist Biosciences, San Francisco, CA, USA), were included in every RT-PCR and sequencing run. Multiplex PCRs were performed in two separate reactions with the different primer pools on a MiniAmp™ Plus Thermal Cycler (Applied Biosystems, Waltham, MA, USA). Each PCR reaction was prepared with half volume of cleaned cDNA (6.25 µL, 8.5 µL and 11.4 µL cleaned cDNA for 150 bp, 400 bp and 1200 bp panels, respectively), 15 nM of each primer, and 1x Q5 Hot Start High-Fidelity Master Mix (New England Biolabs, Ipswich, MA, USA) in a 25 µL reaction volume. The amplification program for the 150 bp amplicons was: heat activation at 98°C for 30 sec, followed by 35 cycles of denaturation at 98 °C for 15 sec and annealing/extension at 65 °C for 2 min. The same amplification program was used for generating 400 bp and 1200 bp amplicons, except that a longer annealing/extension time of 5 min was used.

*Library preparation for sequencing with ONT MinION (400bp and 1200bp amplicons)*

PCR products from the two separate multiplexed reactions for each sample generated with either the 400bp or 1200bp amplicon scheme were pooled and cleaned with 50 µL (1.0x beads/sample ratio) Mag-Bind® TotalPure NGS beads (Omega Bio-tek, Norcross, GA, USA), and quantified with the Qubit™ dsDNA HS Assay Kit (Invitrogen, Waltham, MA, USA). Pooled amplicons were first end-prepared with NEBNext® Ultra™ II End Repair/dA-Tailing Module (New England Biolabs, Ipswich, MA, USA). The end-prepped reactions were composed of 200 fmol amplicons, 1.75 µL End-prep reaction buffer and 0.75 µL Enp-prep enzyme mix in a 15 µL reaction volume. The incubation program was 20°C for 5 min and 65°C for 5 min. Then, barcodes (EXP-NEB104 and EXP-NEB114, Oxford Nanopore Technologies, Oxford, UK) were ligated to end-prepared amplicons with Blunt/TA Ligase Master Mix (New England Biolabs, Ipswich, MA, USA). The barcode ligation reactions were composed of 1.5 µL (20 fmol) end-prepared DNA, 2.5 µL native barcode, and 10 µL Blunt/TA Ligase Master Mix in a 20 µL reaction volume. The incubation program was 20°C for 20 min and 65°C for 10 min. Barcoded samples were pooled and cleaned with Mag-Bind® TotalPure NGS beads (0.4x beads/sample ratio, Omega Bio-tek, Norcross, GA, USA), and quantified with the Qubit™ dsDNA HS Assay Kit (Invitrogen, Waltham, MA, USA). After barcode ligation, sequencing adapters were ligated to barcoded DNAs using Quick T4 DNA Ligase (New England Biolabs, Ipswich, MA, USA). The adapter ligation reactions consisted of 30 µL pooled barcoded library, 5 µL Adapter Mix II (AMII), 5 µL Quick T4 DNA Ligase, and 10 µL NEBNext Quick Ligation reaction buffer in 50 µL reaction volume. The reaction mix was incubated at room temperature for 20 min, cleaned with Mag-Bind® TotalPure NGS beads (0.4x beads/sample ratio, Omega Bio-tek, Norcross, GA, USA), and quantified with the Qubit™ dsDNA HS Assay Kit (Invitrogen, Waltham, MA, USA). Finally, 50 fmol of sequencing library was loaded onto R9.4.1 flowcells using ONT Ligation Sequencing Kit (SQK-LSK109, Oxford Nanopore Technologies, Oxford, UK), and sequenced for 20-48 hours with the MinION device (Oxford Nanopore Technologies, Oxford, UK).

*Library preparation for sequencing with Illumina MiSeq (150bp amplicons)*

PCR products from the two separate multiplexed reactions for each sample generated with the 150bp amplicon scheme were pooled and double-size selected with Mag-Bind® TotalPure NGS beads (0.85x and 1.8x beads/sample ratio, Omega Bio-tek, Norcross, GA, USA), and then quantified with the Qubit™ dsDNA HS Assay Kit (Invitrogen, Waltham, MA, USA). Barcoded sequencing libraries were prepared with 100 ng amplicons using the NEBNext® Ultra™ II DNA Library Prep Kit for Illumina® with NEBNext® Multiplex Oligos for Illumina® (Index Primer Set #1, New England Biolabs, Ipswich, MA, USA) according to the manufacturer's instructions, except that a decreased reaction volume of 24 µL for DNA end-prep reactions and an additional doublesized selection with Mag-Bind® TotalPure NGS beads (0.7x and 1.0x beads/sample ratio) was used before pooling barcoded samples. The pooled library was diluted to 15 mM and sequenced on the Illumina MiSeq v2 2x150bp platform at the UBC Sequencing + Bioinformatics Consortium. A detailed protocol is available at https://www.protocols.io/private/924C2829A15611EB8ED10A58A9FEAC02.

***Bioinformatics analysis***

Illumina raw reads were adapter-trimmed using BBMap v38.86 (3) using BBDuk.sh. Adapter trimmed reads were then quality filtered with Sickle v1.33 (4) using paired end operation and -M option. Nanopore signal data was basecalled and demultiplexed with guppy v4.5.2 using the bonito v3.1 model. Demultiplexed nanopore reads were length and quality filtered using the artic-ncov2019 pipeline according to the suggested parameters (https://artic.network/ncov-2019/ncov2019-bioinformatics-sop.html). Filtered reads were mapped to the SARS-CoV-2 Wuhan-Hu-1 reference genome sequence (NCBI accession MN908947.3) using minimap2 v.2.17 (5) using the option -ax sr (Illumina) or -ax map-ont (nanopore). Mapping files were parsed using samtools v.1.12 (6) to determine the frequency of variant of concern (VoC)-associated SNVs.

VoC-associated SNVs were determined by mapping sequences from the P.1 (n=2,170), B.1.1.7 (n=440,207), and B.1.351 (n=11,809) lineages available from GISAID (https://www.gisaid.org, downloaded April 23, 2021) against the Wuhan-Hu-1 reference genome sequence using minimap2 v.2.17 with the option -ax asm5. SNVs detected above 90% frequency were then queried for their occurrence in other SARS-CoV-2 lineages by comparison to all available SARS-CoV-2 genome sequences in GISAID (n=1,194,352, on April 23, 2021) using BAMQL v.1.6 (7). SNVs present at over 10% frequency in another lineage were then filtered from the VoC-associated SNV lists, yielding a total of 26, 17, and 11 lineage-specific SNVs for B.1.1.7, P.1, and B.1.351, respectively (Table S3).

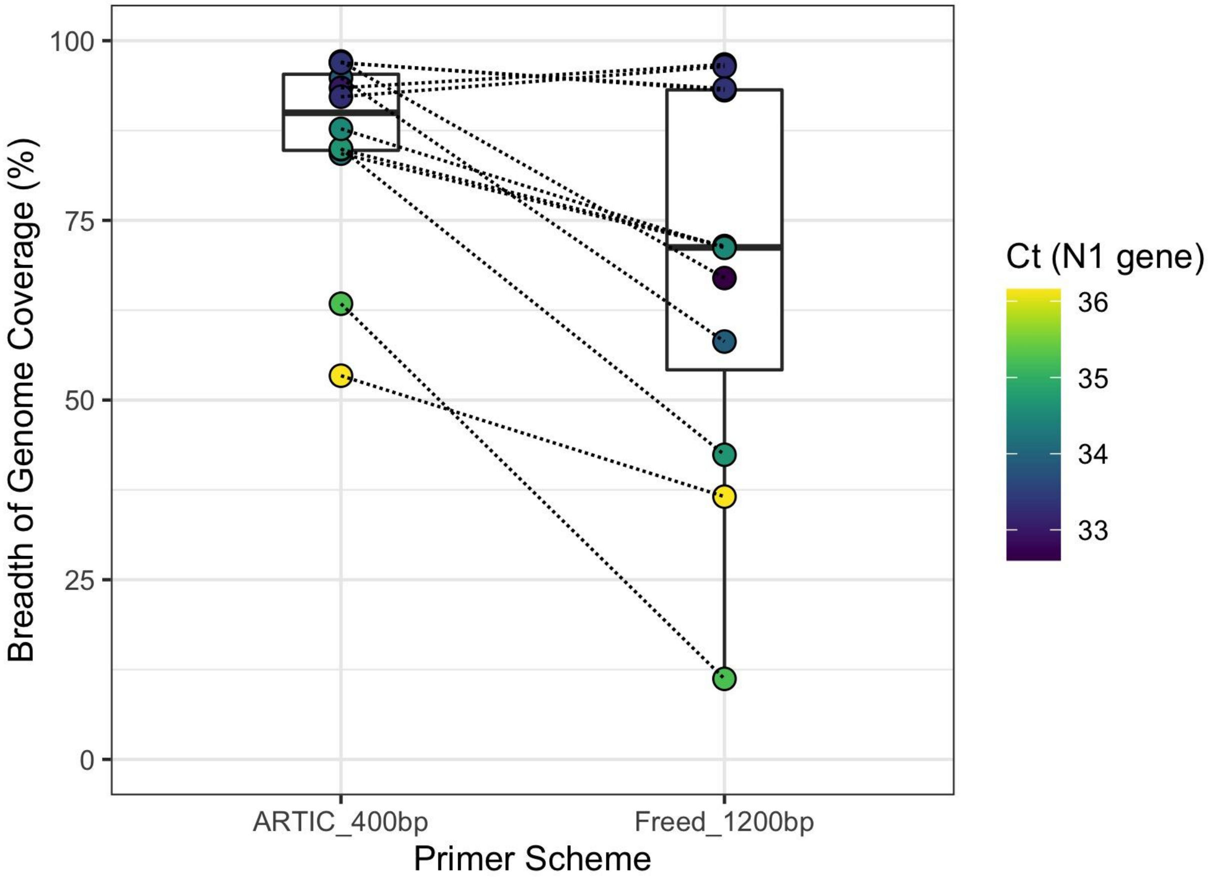


**Figure S1.** Breadth of genome coverage for paired samples with $C_t$ values more than 32.5 sequenced with both the 400bp and 1200bp primer schemes. Points are colored based on the sample $C_t$ value (US CDC N1 gene, see Text S1), and the dotted line indicates a sample pair.

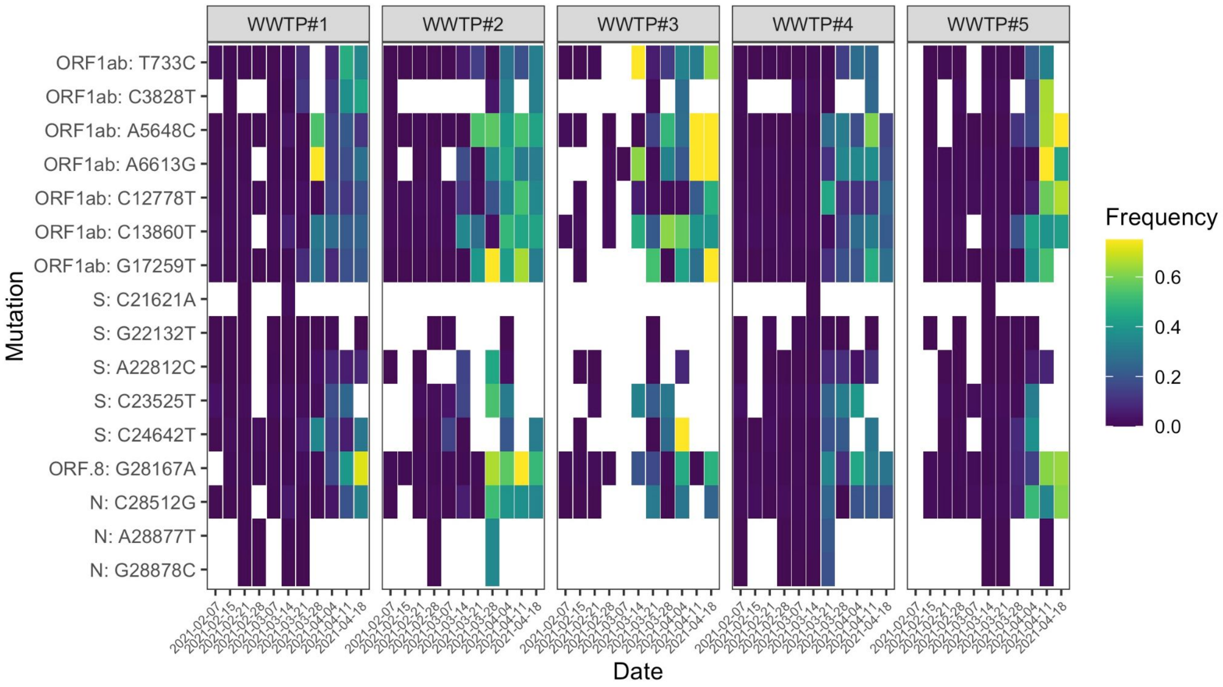


**Figure S2.** Heatmap showing the frequencies of single nucleotide variants (SNVs) associated with the P.1 lineage over the study period in the influent wastewater of five municipal WWTPs in Metro Vancouver, British Columbia. Values are shown only for SNVs detected with a total read coverage of 50 or greater at that genomic position, and otherwise the value is shown as white. SNVs that never had a read coverage over 50 across all samples were filtered. All P.1-associated SNV sites that were queried are provided in Table S3.

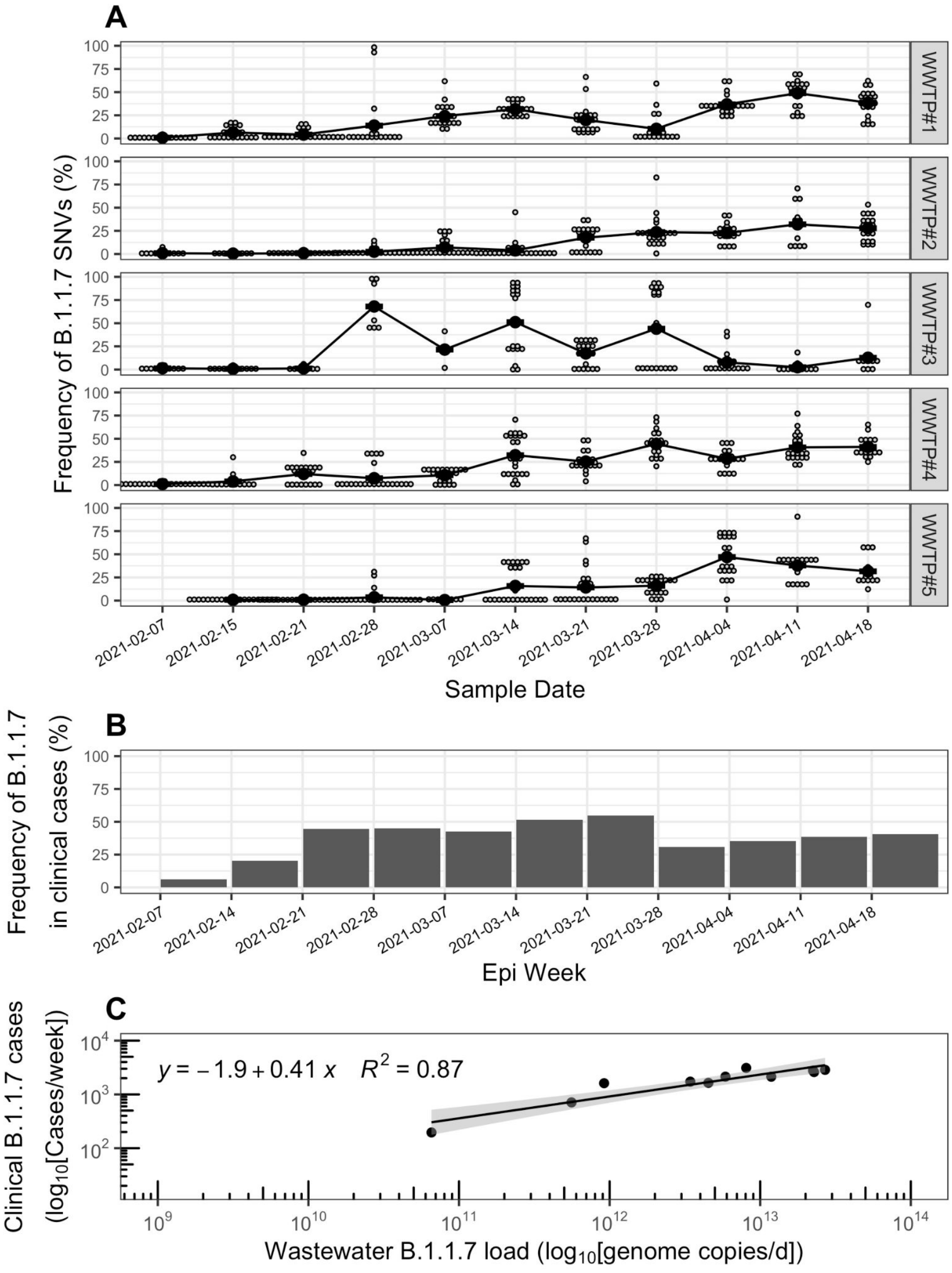


**Figure S3. (A)** Frequency of SNVs associated with the B.1.1.7 lineage of SARS-CoV-2 within influent wastewater samples from five wastewater treatment plants in Metro Vancouver, British Columbia (BC), from February 7th to April 18th, 2021. Smaller grey dots represent the frequency of individual SNVs, while the larger black points represent the mean frequency across all detected SNVs. Only genome positions with a read coverage over 50 are included in SNV frequency calculations. **(B)** Frequency of the B.1.1.7 lineage in clinical COVID-19 patient cases in the province of BC, Canada over the study period. The frequencies in clinical patient cases correspond to an average value detected over an epidemiology (epi) week, and were adapted from (8). **(C)** Correlation between the wastewater cumulative daily load of B.1.1.7 genomes summed across all five WWTPs and the total B.1.1.7 clinical cases in the province of BC observed within the same epidemiological week. The wastewater B.1.1.7 daily load (genome copies/d) was approximated by normalizing copies of the N1 gene (copies/L) by daily flow rates (L/d) to obtain N1 loads (copies/d) for all WWTPs, and multiplying that by the mean frequency of B.1.1.7-associated SNVs in each WWTP across all sample dates. For each date, the cumulative B.1.1.7 daily load was determined by summing the B.1.1.7 loads across all five WWTPs. The B.1.1.7 clinical case counts were adapted from (8) by multiplying total provincial COVID-19 case counts by the frequency of B.1.1.7 in clinical provincial cases. Data points with zero clinical cases are shown aligned to the x-axis.

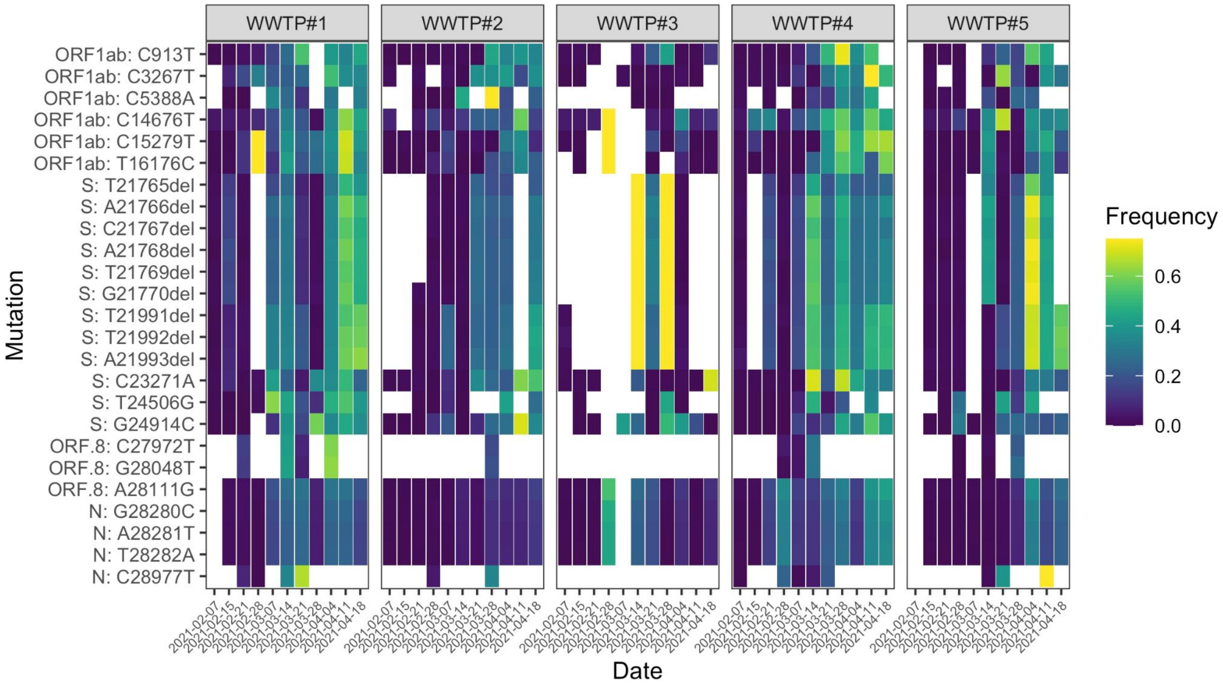


**Figure S4.** Heatmap showing the frequencies of SNVs associated with the B.1.1.7 lineage over the study period in the influent wastewater five municipal WWTPs in Metro Vancouver, British Columbia. Values are shown only for SNVs detected with a total read coverage of 50 or greater at that genomic position, and otherwise the value is shown as white. SNVs that never had a read coverage over 50 across all samples were filtered. All B.1.1.7-associated SNV sites that were queried are provided in Table S3.

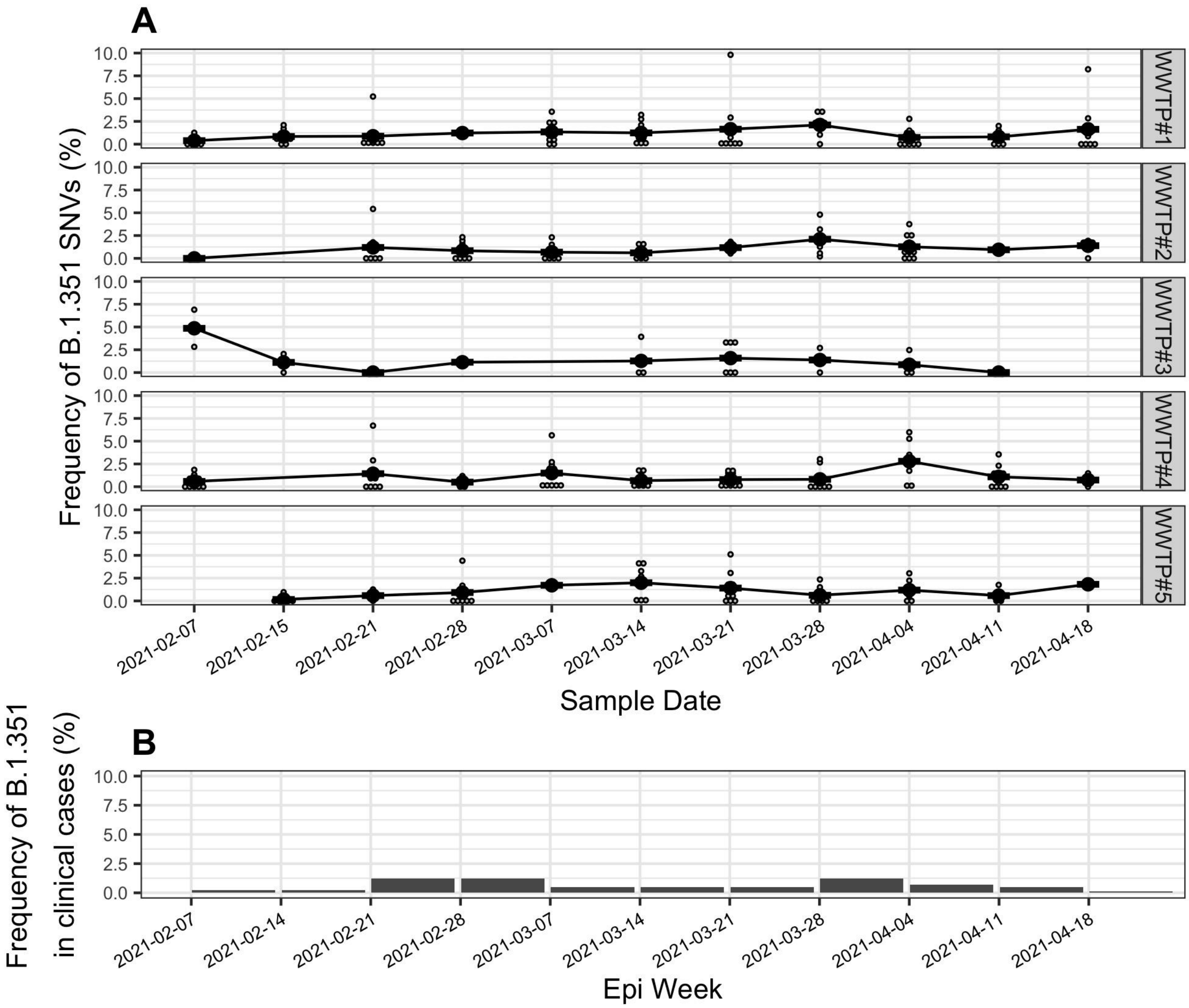


**Figure S5. (A)** Frequency of SNVs associated with the B.1.351 lineage of SARS-CoV-2 within influent wastewater samples from five wastewater treatment plants in Metro Vancouver, British Columbia (BC), from February 7th to April 18th, 2021. Smaller grey dots represent the frequency of individual SNVs, while the larger black points represent the mean across all detected SNVs. Only genome positions with a read coverage over 50 are included in SNV frequency calculations. **(B)** Frequency of the B.1.351 lineage in clinical COVID-19 patient cases in the province of BC, Canada over the study period. The frequencies in clinical patient cases correspond to an average value detected over an epidemiology (epi) week, and were adapted from (8).

**Table S1:** Sample metadata, including sample epidemiological week and sampling date, WWTP, sample type, daily average flow rate, $C_t$ of the US CDC N1 assay, sequencing library type, sequencing throughput, mapping rates, and NCBI accession for raw read data. For sample type, WW indicates influent wastewater and PS indicates primary sludge.

| Epi Week | Date sampled | WWTP | Sample type | Flow rate (MLD) | US CDC N1 RT-qPCR | | Amplicon size (bp) | Amplicon concentration (post PCR) (ng/µL) | Sequencing Platform | Raw bases (bp) | Bases mapped (bp) | Mapping rate | NCBI accession number |
|---|---|---|---|---|---|---|---|---|---|---|---|---|---|
| | | | | | Ct | Quantity (copies/L influent or copies/g-TS sludge) | | | | | | | |
| 5 | 07-Feb-21 | WWTP#1 | WW | 596 | 33.66 | 6.81E+03 | 400 | 13.2 | ONT_MinION | 8.35E+08 | 1.86E+07 | 2.2% | SAMN19321363 |
| 5 | 07-Feb-21 | WWTP#1 | PS | N/A | 31.78 | N/A | 400 | 25.4 | ONT_MinION | 4.78E+07 | 7.61E+02 | 0.0% | SAMN19321367 |
| 5 | 07-Feb-21 | WWTP#2 | WW | 527 | 34.20 | 4.67E+03 | 400 | 28.0 | ONT_MinION | 1.09E+08 | 1.19E+07 | 10.9% | SAMN19321364 |
| 5 | 07-Feb-21 | WWTP#3 | WW | 90 | 34.54 | 3.74E+03 | 400 | 54.4 | ONT_MinION | 1.62E+08 | 3.62E+06 | 2.2% | SAMN19304050 |
| 5 | 07-Feb-21 | WWTP#3 | WW | 90 | 34.54 | 3.74E+03 | 1200 | 26.6 | ONT_MinION | 4.33E+08 | 4.43E+06 | 1.0% | SAMN19304105 |
| 5 | 07-Feb-21 | WWTP#4 | WW | 72 | 33.61 | 6.97E+03 | 400 | 22.0 | ONT_MinION | 5.47E+08 | 5.34E+07 | 9.7% | SAMN19321365 |
| 6 | 15-Feb-21 | WWTP#1 | WW | 593 | 32.60 | 1.38E+04 | 400 | 52.4 | ONT_MinION | 3.63E+08 | 3.11E+07 | 8.6% | SAMN19304051 |
| 6 | 15-Feb-21 | WWTP#1 | PS | N/A | 30.78 | N/A | 400 | 27.0 | ONT_MinION | 1.25E+08 | 6.45E+05 | 0.5% | SAMN19321368 |
| 6 | 15-Feb-21 | WWTP#1 | WW | 593 | 32.60 | 1.38E+04 | 1200 | 26.8 | ONT_MinION | 6.47E+08 | 3.86E+06 | 0.6% | SAMN19304106 |
| 6 | 15-Feb-21 | WWTP#2 | WW | 997 | 34.28 | 4.38E+03 | 150 | 27.2 | Illumina_MiSeq | 4.51E+08 | 7.10E+06 | 1.6% | SAMN19304040 |
| 6 | 15-Feb-21 | WWTP#2 | WW | 997 | 34.28 | 4.38E+03 | 400 | 55.8 | ONT_MinION | 6.63E+07 | 3.26E+06 | 4.9% | SAMN19304052 |
| 6 | 15-Feb-21 | WWTP#2 | WW | 997 | 34.28 | 4.38E+03 | 1200 | 28.2 | ONT_MinION | 4.02E+08 | 8.30E+06 | 2.1% | SAMN19304107 |
| 6 | 15-Feb-21 | WWTP#3 | WW | 88 | 35.67 | 1.84E+03 | 400 | 26.4 | ONT_MinION | 2.12E+08 | 1.39E+07 | 6.5% | SAMN19321366 |
| 6 | 15-Feb-21 | WWTP#4 | WW | 76 | 34.41 | 4.23E+03 | 400 | 53.6 | ONT_MinION | 2.71E+08 | 1.10E+07 | 4.1% | SAMN19304053 |
| 6 | 15-Feb-21 | WWTP#5 | WW | 15 | 34.80 | 3.14E+03 | 400 | 50.8 | ONT_MinION | 3.68E+08 | 9.32E+06 | 2.5% | SAMN19304054 |
| 7 | 21-Feb-21 | WWTP#1 | WW | 600 | 31.75 | 2.73E+04 | 150 | 27.4 | Illumina_MiSeq | 4.63E+08 | 2.38E+07 | 5.2% | SAMN19304041 |
| 7 | 21-Feb-21 | WWTP#1 | WW | 600 | 31.75 | 2.73E+04 | 400 | 38.2 | ONT_MinION | 3.80E+08 | 1.50E+07 | 4.0% | SAMN19304055 |
| 7 | 21-Feb-21 | WWTP#1 | PS | N/A | 33.06 | N/A | 400 | 23.2 | ONT_MinION | 6.45E+07 | 3.92E+03 | 0.0% | SAMN19321369 |
| 7 | 21-Feb-21 | WWTP#2 | WW | 626 | 32.64 | 1.50E+04 | 400 | 48.8 | ONT_MinION | 9.27E+07 | 2.80E+06 | 3.0% | SAMN19304056 |
| 7 | 21-Feb-21 | WWTP#3 | WW | 93 | 35.24 | 2.89E+03 | 400 | 44.6 | ONT_MinION | 1.33E+08 | 1.31E+06 | 1.0% | SAMN19304057 |

| | | | | | | | | | | | | | |
|---|---|---|---|---|---|---|---|---|---|---|---|---|---|
| 7 | 21-Feb-21 | WWTP#3 | WW | 93 | 35.24 | 2.89E+03 | 1200 | 21.0 | ONT_MinION | 3.93E+08 | 1.06E+06 | 0.3% | SAMN19304108 |
| 7 | 21-Feb-21 | WWTP#4 | WW | 71 | 32.43 | 1.72E+04 | 400 | 40.0 | ONT_MinION | 3.68E+08 | 1.04E+07 | 2.8% | SAMN19304058 |
| 7 | 21-Feb-21 | WWTP#5 | WW | 17 | 33.90 | 7.09E+03 | 400 | 39.2 | ONT_MinION | 4.29E+08 | 5.79E+06 | 1.4% | SAMN19304059 |
| 7 | 21-Feb-21 | WWTP#5 | WW | 17 | 33.90 | 7.09E+03 | 1200 | 19.8 | ONT_MinION | 4.32E+08 | 1.45E+07 | 3.3% | SAMN19304109 |
| 8 | 28-Feb-21 | WWTP#1 | PS | N/A | 30.57 | 1.32E+05 | 150 | 25.0 | Illumina_MiSeq | 3.74E+08 | 8.77E+05 | 0.2% | SAMN19304042 |
| 8 | 28-Feb-21 | WWTP#1 | WW | 565 | 32.14 | 3.41E+04 | 400 | 10.3 | ONT_MinION | 3.65E+08 | 7.82E+06 | 2.1% | SAMN19304061 |
| 8 | 28-Feb-21 | WWTP#1 | PS | N/A | 30.57 | 1.32E+05 | 400 | 29.8 | ONT_MinION | 1.58E+08 | 6.68E+05 | 0.4% | SAMN19304060 |
| 8 | 28-Feb-21 | WWTP#1 | WW | 565 | 32.14 | 3.41E+04 | 1200 | 16.0 | ONT_MinION | 7.15E+08 | 5.27E+07 | 7.4% | SAMN19304111 |
| 8 | 28-Feb-21 | WWTP#1 | PS | N/A | 30.57 | 1.32E+05 | 1200 | 18.7 | ONT_MinION | 3.41E+08 | 4.06E+02 | 0.0% | SAMN19304110 |
| 8 | 28-Feb-21 | WWTP#2 | WW | 639 | 33.25 | 1.80E+04 | 400 | 29.6 | ONT_MinION | 3.43E+08 | 9.00E+06 | 2.6% | SAMN19304062 |
| 8 | 28-Feb-21 | WWTP#3 | WW | 83 | 34.58 | 6.59E+03 | 400 | 36.6 | ONT_MinION | 3.11E+08 | 8.28E+05 | 0.3% | SAMN19304063 |
| 8 | 28-Feb-21 | WWTP#4 | WW | 70 | 33.38 | 1.53E+04 | 400 | 33.2 | ONT_MinION | 4.10E+08 | 3.25E+06 | 0.8% | SAMN19304064 |
| 8 | 28-Feb-21 | WWTP#5 | WW | 16 | 32.64 | 2.44E+04 | 400 | 34.4 | ONT_MinION | 4.74E+08 | 1.11E+07 | 2.4% | SAMN19304065 |
| 9 | 07-Mar-21 | WWTP#1 | PS | N/A | 30.78 | 8.22E+04 | 150 | 30.8 | Illumina_MiSeq | 4.08E+08 | 3.01E+05 | 0.1% | SAMN19304043 |
| 9 | 07-Mar-21 | WWTP#1 | WW | 555 | 32.40 | 2.85E+04 | 400 | 38.8 | ONT_MinION | 3.65E+08 | 6.10E+07 | 16.7% | SAMN19304068 |
| 9 | 07-Mar-21 | WWTP#1 | PS | N/A | 30.78 | 8.22E+04 | 400 | 38.0 | ONT_MinION | 1.72E+08 | 5.48E+04 | 0.0% | SAMN19304066 |
| 9 | 07-Mar-21 | WWTP#1 | WW | 555 | 32.40 | 2.85E+04 | 1200 | 30.6 | ONT_MinION | 4.91E+08 | 7.92E+07 | 16.1% | SAMN19304114 |
| 9 | 07-Mar-21 | WWTP#1 | PS | N/A | 30.78 | 8.22E+04 | 1200 | 17.4 | ONT_MinION | 3.94E+08 | 0.00E+00 | 0.0% | SAMN19304112 |
| 9 | 07-Mar-21 | WWTP#2 | WW | 532 | 33.39 | 1.47E+04 | 150 | 33.4 | Illumina_MiSeq | 4.35E+08 | 1.22E+07 | 2.8% | SAMN19304044 |
| 9 | 07-Mar-21 | WWTP#2 | WW | 532 | 33.39 | 1.47E+04 | 400 | 51.2 | ONT_MinION | 2.34E+08 | 2.05E+07 | 8.8% | SAMN19304069 |
| 9 | 07-Mar-21 | WWTP#2 | PS | N/A | 31.39 | 5.32E+04 | 400 | 25.0 | ONT_MinION | 2.02E+08 | 4.86E+04 | 0.0% | SAMN19304067 |
| 9 | 07-Mar-21 | WWTP#2 | WW | 532 | 33.39 | 1.47E+04 | 1200 | 25.4 | ONT_MinION | 5.21E+08 | 4.56E+07 | 8.7% | SAMN19304115 |
| 9 | 07-Mar-21 | WWTP#2 | PS | N/A | 31.39 | 5.32E+04 | 1200 | 13.4 | ONT_MinION | 2.48E+08 | 1.15E+04 | 0.0% | SAMN19304113 |
| 9 | 07-Mar-21 | WWTP#3 | WW | 87 | 36.16 | 2.52E+03 | 150 | 32.0 | Illumina_MiSeq | 1.48E+08 | 5.62E+05 | 0.4% | SAMN19304045 |
| 9 | 07-Mar-21 | WWTP#3 | WW | 87 | 36.16 | 2.52E+03 | 400 | 45.4 | ONT_MinION | 2.44E+08 | 1.69E+06 | 0.7% | SAMN19304070 |
| 9 | 07-Mar-21 | WWTP#3 | WW | 87 | 36.16 | 2.52E+03 | 1200 | 33.4 | ONT_MinION | 3.98E+08 | 2.61E+06 | 0.7% | SAMN19304116 |
| 9 | 07-Mar-21 | WWTP#4 | WW | 69 | 33.35 | 1.51E+04 | 400 | 35.2 | ONT_MinION | 3.52E+08 | 2.09E+07 | 5.9% | SAMN19304071 |

| | | | | | | | | | | | | | |
|---|---|---|---|---|---|---|---|---|---|---|---|---|---|
| 9 | 07-Mar-21 | WWTP#4 | WW | 69 | 33.35 | 1.51E+04 | 1200 | 19.7 | ONT_MinION | 6.26E+08 | 5.80E+07 | 9.3% | SAMN19304117 |
| 9 | 07-Mar-21 | WWTP#5 | WW | 16 | 36.40 | 1.89E+03 | 150 | 25.6 | Illumina_MiSeq | 3.77E+08 | 5.04E+06 | 1.3% | SAMN19304046 |
| 9 | 07-Mar-21 | WWTP#5 | WW | 16 | 36.40 | 1.89E+03 | 400 | 32.0 | ONT_MinION | 4.48E+08 | 8.64E+06 | 1.9% | SAMN19304072 |
| 10 | 14-Mar-21 | WWTP#1 | PS | N/A | 32.53 | 4.11E+04 | 150 | 22.2 | Illumina_MiSeq | 3.69E+08 | 3.03E+05 | 0.1% | SAMN19304047 |
| 10 | 14-Mar-21 | WWTP#1 | WW | 488 | 31.15 | 6.67E+04 | 400 | 46.6 | ONT_MinION | 4.02E+08 | 1.10E+08 | 27.5% | SAMN19304075 |
| 10 | 14-Mar-21 | WWTP#1 | PS | N/A | 32.53 | 4.11E+04 | 400 | 36.6 | ONT_MinION | 1.60E+08 | 4.31E+04 | 0.0% | SAMN19304073 |
| 10 | 14-Mar-21 | WWTP#1 | WW | 488 | 31.15 | 6.67E+04 | 1200 | 26.6 | ONT_MinION | 7.30E+08 | 9.31E+07 | 12.8% | SAMN19304119 |
| 10 | 14-Mar-21 | WWTP#1 | PS | N/A | 32.53 | 4.11E+04 | 1200 | 19.9 | ONT_MinION | 2.95E+08 | 1.56E+04 | 0.0% | SAMN19304118 |
| 10 | 14-Mar-21 | WWTP#2 | WW | 444 | 32.97 | 1.98E+04 | 150 | 22.2 | Illumina_MiSeq | 3.90E+08 | 1.31E+07 | 3.4% | SAMN19304048 |
| 10 | 14-Mar-21 | WWTP#2 | WW | 444 | 32.97 | 1.98E+04 | 400 | 33.2 | ONT_MinION | 1.18E+08 | 1.61E+07 | 13.6% | SAMN19304076 |
| 10 | 14-Mar-21 | WWTP#2 | PS | N/A | 28.92 | 3.44E+05 | 400 | 42.4 | ONT_MinION | 2.78E+08 | 1.70E+05 | 0.1% | SAMN19304074 |
| 10 | 14-Mar-21 | WWTP#2 | WW | 444 | 32.97 | 1.98E+04 | 1200 | 17.9 | ONT_MinION | 8.00E+08 | 5.17E+07 | 6.5% | SAMN19304120 |
| 10 | 14-Mar-21 | WWTP#3 | WW | 74 | 34.66 | 6.21E+03 | 400 | 38.0 | ONT_MinION | 1.95E+08 | 8.80E+06 | 4.5% | SAMN19304077 |
| 10 | 14-Mar-21 | WWTP#3 | WW | 74 | 34.66 | 6.21E+03 | 1200 | 17.1 | ONT_MinION | 8.16E+08 | 1.06E+07 | 1.3% | SAMN19304121 |
| 10 | 14-Mar-21 | WWTP#4 | WW | 69 | 31.87 | 4.19E+04 | 400 | 35.8 | ONT_MinION | 4.18E+08 | 8.08E+07 | 19.3% | SAMN19304078 |
| 10 | 14-Mar-21 | WWTP#5 | WW | 15 | 32.79 | 2.33E+04 | 400 | 32.6 | ONT_MinION | 4.45E+08 | 5.79E+07 | 13.0% | SAMN19304079 |
| 11 | 21-Mar-21 | WWTP#1 | WW | 544 | 31.63 | 4.81E+04 | 400 | 32.2 | ONT_MinION | 3.08E+08 | 1.12E+08 | 36.5% | SAMN19304080 |
| 11 | 21-Mar-21 | WWTP#1 | WW | 544 | 31.63 | 4.81E+04 | 1200 | 23.0 | ONT_MinION | 4.88E+08 | 5.97E+07 | 12.2% | SAMN19304122 |
| 11 | 21-Mar-21 | WWTP#2 | WW | 686 | 33.29 | 1.59E+04 | 150 | 39.2 | Illumina_MiSeq | 4.46E+08 | 2.09E+07 | 4.7% | SAMN19304049 |
| 11 | 21-Mar-21 | WWTP#2 | WW | 686 | 33.29 | 1.59E+04 | 400 | 39.4 | ONT_MinION | 7.45E+07 | 2.43E+07 | 32.6% | SAMN19304081 |
| 11 | 21-Mar-21 | WWTP#2 | WW | 686 | 33.29 | 1.59E+04 | 1200 | 18.7 | ONT_MinION | 4.77E+08 | 5.21E+07 | 10.9% | SAMN19304123 |
| 11 | 21-Mar-21 | WWTP#3 | WW | 95 | 33.31 | 1.59E+04 | 400 | 43.4 | ONT_MinION | 1.40E+08 | 3.65E+07 | 26.0% | SAMN19304082 |
| 11 | 21-Mar-21 | WWTP#4 | WW | 66 | 31.99 | 3.97E+04 | 400 | 32.0 | ONT_MinION | 3.35E+08 | 9.36E+07 | 27.9% | SAMN19304083 |
| 11 | 21-Mar-21 | WWTP#4 | WW | 66 | 31.99 | 3.97E+04 | 1200 | 17.3 | ONT_MinION | 5.27E+08 | 5.31E+07 | 10.1% | SAMN19304124 |
| 11 | 21-Mar-21 | WWTP#5 | WW | 16 | 34.39 | 9.00E+03 | 400 | 30.0 | ONT_MinION | 3.43E+08 | 8.08E+07 | 23.6% | SAMN19304084 |
| 12 | 28-Mar-21 | WWTP#1 | WW | 584 | 32.09 | 3.56E+04 | 400 | 41.2 | ONT_MinION | 1.65E+08 | 2.41E+07 | 14.7% | SAMN19304085 |
| 12 | 28-Mar-21 | WWTP#1 | WW | 584 | 32.09 | 3.56E+04 | 1200 | 34.6 | ONT_MinION | 4.44E+08 | 1.82E+07 | 4.1% | SAMN19304125 |

| | | | | | | | | | | | | | |
|---|---|---|---|---|---|---|---|---|---|---|---|---|---|
| 12 | 28-Mar-21 | WWTP#2 | WW | 566 | 33.03 | 1.86E+04 | 400 | 35.2 | ONT_MinION | 3.21E+08 | 3.06E+07 | 9.5% | SAMN19304086 |
| 12 | 28-Mar-21 | WWTP#3 | WW | 86 | 34.50 | 7.40E+03 | 400 | 50.6 | ONT_MinION | 1.93E+08 | 1.65E+07 | 8.5% | SAMN19304087 |
| 12 | 28-Mar-21 | WWTP#3 | WW | 86 | 34.50 | 7.40E+03 | 1200 | 23.8 | ONT_MinION | 5.28E+08 | 1.89E+07 | 3.6% | SAMN19304126 |
| 12 | 28-Mar-21 | WWTP#4 | WW | 65 | 32.28 | 3.09E+04 | 400 | 43.0 | ONT_MinION | 1.12E+08 | 1.97E+07 | 17.7% | SAMN19304088 |
| 12 | 28-Mar-21 | WWTP#5 | WW | 16 | 33.45 | 1.53E+04 | 400 | 47.8 | ONT_MinION | 1.15E+08 | 2.17E+07 | 18.8% | SAMN19304089 |
| 13 | 04-Apr-21 | WWTP#1 | WW | 469 | 30.34 | 1.15E+05 | 400 | 43.6 | ONT_MinION | 2.02E+08 | 7.42E+07 | 36.6% | SAMN19304090 |
| 13 | 04-Apr-21 | WWTP#1 | WW | 469 | 30.34 | 1.15E+05 | 1200 | 24.4 | ONT_MinION | 4.20E+08 | 7.70E+07 | 18.3% | SAMN19304127 |
| 13 | 04-Apr-21 | WWTP#2 | WW | 370 | 31.20 | 6.41E+04 | 400 | 44.8 | ONT_MinION | 2.21E+08 | 6.54E+07 | 29.5% | SAMN19304091 |
| 13 | 04-Apr-21 | WWTP#2 | WW | 370 | 31.20 | 6.41E+04 | 1200 | 28.0 | ONT_MinION | 2.90E+08 | 3.04E+07 | 10.5% | SAMN19304128 |
| 13 | 04-Apr-21 | WWTP#3 | WW | 69 | 34.30 | 7.89E+03 | 400 | 26.8 | ONT_MinION | 1.48E+08 | 3.68E+07 | 24.9% | SAMN19304092 |
| 13 | 04-Apr-21 | WWTP#4 | WW | 64 | 30.72 | 8.90E+04 | 400 | 35.8 | ONT_MinION | 6.31E+07 | 1.82E+07 | 28.8% | SAMN19304093 |
| 13 | 04-Apr-21 | WWTP#5 | WW | 14 | 33.81 | 1.13E+04 | 400 | 25.6 | ONT_MinION | 1.19E+08 | 3.10E+07 | 26.1% | SAMN19304094 |
| 14 | 11-Apr-21 | WWTP#1 | WW | 463 | 31.13 | 6.84E+04 | 400 | 39.4 | ONT_MinION | 3.66E+08 | 8.11E+07 | 22.2% | SAMN19304095 |
| 14 | 11-Apr-21 | WWTP#2 | WW | 364 | 31.82 | 4.23E+04 | 400 | 41.4 | ONT_MinION | 7.10E+07 | 1.96E+07 | 27.6% | SAMN19304096 |
| 14 | 11-Apr-21 | WWTP#3 | WW | 72 | 33.29 | 1.64E+04 | 400 | 32.8 | ONT_MinION | 9.78E+07 | 2.68E+07 | 27.4% | SAMN19304097 |
| 14 | 11-Apr-21 | WWTP#4 | WW | 64 | 31.06 | 7.21E+04 | 400 | 33.8 | ONT_MinION | 3.51E+08 | 9.48E+07 | 27.0% | SAMN19304098 |
| 14 | 11-Apr-21 | WWTP#5 | WW | 14 | 32.55 | 2.58E+04 | 400 | 38.8 | ONT_MinION | 2.91E+08 | 6.91E+07 | 23.7% | SAMN19304099 |
| 15 | 18-Apr-21 | WWTP#1 | WW | 452 | 30.92 | 7.86E+04 | 400 | 41.8 | ONT_MinION | 1.34E+08 | 2.71E+07 | 20.2% | SAMN19304100 |
| 15 | 18-Apr-21 | WWTP#2 | WW | 352 | 31.05 | 7.19E+04 | 400 | 38.2 | ONT_MinION | 1.50E+08 | 2.03E+07 | 13.6% | SAMN19304101 |
| 15 | 18-Apr-21 | WWTP#3 | WW | 67 | 34.61 | 7.20E+03 | 400 | 47.2 | ONT_MinION | 1.11E+08 | 7.30E+06 | 6.5% | SAMN19304102 |
| 15 | 18-Apr-21 | WWTP#4 | WW | 68 | 31.14 | 6.69E+04 | 400 | 51.4 | ONT_MinION | 4.11E+07 | 6.62E+06 | 16.1% | SAMN19304103 |
| 15 | 18-Apr-21 | WWTP#5 | WW | 14 | 33.05 | 1.85E+04 | 400 | 40.6 | ONT_MinION | 4.11E+07 | 5.20E+06 | 12.7% | SAMN19304104 |

**Table S2:** Results of RT-PCR inhibition test with Veto VetMax Internal Positive Control (IPC) Assay and BSA-amended US CDC N1 assay. SARS-CoV-2 viral concentrations in primary sludge samples were quantified using the Reliance One-Step Multiplex RT-qPCR Mastermix (Bio-Rad Laboratories, Hercules, CA, USA) with the 2019-nCoV CDC RUO primers and probes targeting the N1 gene (Integrated DNA Technologies). RT-qPCR reactions were prepared as described by D'Aoust et al. (9), and run in triplicate on a CFX96 Real-time System (Bio-Rad Laboratories).

| Sample | VetMax IPC | | US CDC N1 (N1 only) | | US CDC N1 (with IPC) | | US CDC N1 (with BSA) | |
|---|---|---|---|---|---|---|---|---|
| | Lower CI* | Upper CI* | Ct | Ct Stdev. | Ct | Ct Stdev. | Ct | Ct Stdev. |
| Wk5 WWTP#1 | 30.16 | 30.57 | 34.31 | 0.26 | 34.77 | 0.45 | 34.64 | 0.42 |
| Wk6 WWTP#1 | 29.91 | 30.92 | 36.72 | 0.16 | 35.67 | 0.25 | 35.44 | 1.27 |
| Wk7 WWTP#1 | 30.08 | 30.95 | 36.38 | 0.79 | 36.59 | 0.78 | 36.01 | 0.56 |
| Wk8 WWTP#1 | 30.35 | 31.38 | 33.79 | 0.20 | 34.12 | 0.52 | 33.92 | 0.44 |
| Wk9 WWTP#1 | 30.11 | 30.96 | 33.57 | 0.27 | 33.34 | 0.27 | 33.24 | 0.09 |
| Wk10 WWTP#1 | 30.08 | 30.70 | 34.60 | 0.49 | 35.11 | 0.68 | 34.44 | 0.30 |
| Wk9 WWTP#2 | 30.26 | 30.74 | 34.30 | 0.10 | 34.37 | 0.36 | 34.19 | 0.27 |
| Wk10 WWTP#2 | 30.16 | 30.78 | 33.67 | 0.30 | 33.71 | 0.37 | 33.73 | 0.66 |
| PC | 30.96 | 31.43 | 33.26 | 0.44 | 33.07 | 0.25 | 33.13 | 0.80 |
| NTC | 30.55 | 32.39 | NA | 0.00 | NA | 0.00 | 43.44 | 0.00** |

* $C_t$ values were calculated for 95% confidence intervals
** Detected in only one of the three technical replicates

**Table S3:** Single nucleotide variants (SNVs) associated with SARS-CoV-2 lineages of concern (or variants of concern). The mutations were identified by querying the GISAID database to find SNVs that were over 90% prevalent in the variants of concern, and were also not present above 10% prevalence in another lineage (see Text S1).

| Lineage | Nucleotide Mutation | Gene | Prevalence in lineage (in GISAID) |
|---|---|---|---|
| P.1 | T733C | ORF1ab | 0.999 |
| P.1 | C2749T | ORF1ab | 0.991 |
| P.1 | C3828T | ORF1ab | 0.993 |
| P.1 | A5648C | ORF1ab | 0.998 |
| P.1 | A6613G | ORF1ab | 0.995 |
| P.1 | C12778T | ORF1ab | 0.988 |
| P.1 | C13860T | ORF1ab | 0.992 |
| P.1 | G17259T | ORF1ab | 0.996 |
| P.1 | C21621A | S | 0.994 |
| P.1 | G22132T | S | 0.972 |
| P.1 | A22812C | S | 0.913 |
| P.1 | C23525T | S | 0.998 |
| P.1 | C24642T | S | 0.996 |
| P.1 | G28167A | ORF.8 | 0.972 |
| P.1 | C28512G | N | 0.992 |
| P.1 | A28877T | N | 0.945 |
| P.1 | G28878C | N | 0.930 |
| B.1.1.7 | C913T | ORF1ab | 0.989 |
| B.1.1.7 | C3267T | ORF1ab | 0.998 |
| B.1.1.7 | C5388A | ORF1ab | 0.997 |
| B.1.1.7 | T6954C | ORF1ab | 0.988 |
| B.1.1.7 | C14676T | ORF1ab | 0.998 |
| B.1.1.7 | C15279T | ORF1ab | 0.997 |
| B.1.1.7 | T16176C | ORF1ab | 0.998 |
| B.1.1.7 | T21765del | S | 0.974 |
| B.1.1.7 | A21766del | S | 0.975 |
| B.1.1.7 | C21767del | S | 0.978 |
| B.1.1.7 | A21768del | S | 0.978 |
| B.1.1.7 | T21769del | S | 0.978 |

| | | | |
|---|---|---|---|
| B.1.1.7 | G21770del | S | 0.978 |
| B.1.1.7 | T21991del | S | 0.970 |
| B.1.1.7 | T21992del | S | 0.969 |
| B.1.1.7 | A21993del | S | 0.969 |
| B.1.1.7 | C23271A | S | 0.996 |
| B.1.1.7 | T24506G | S | 0.998 |
| B.1.1.7 | G24914C | S | 0.997 |
| B.1.1.7 | C27972T | ORF.8 | 0.998 |
| B.1.1.7 | G28048T | ORF.8 | 0.995 |
| B.1.1.7 | A28111G | ORF.8 | 0.998 |
| B.1.1.7 | G28280C | N | 0.988 |
| B.1.1.7 | A28281T | N | 0.988 |
| B.1.1.7 | T28282A | N | 0.990 |
| B.1.1.7 | C28977T | N | 0.996 |
| B.1.351 | G5230T | ORF1ab | 0.991 |
| B.1.351 | A10323G | ORF1ab | 0.996 |
| B.1.351 | A21801C | S | 0.996 |
| B.1.351 | A22206G | S | 0.966 |
| B.1.351 | T22287del | S | 0.925 |
| B.1.351 | T22288del | S | 0.925 |
| B.1.351 | G22289del | S | 0.923 |
| B.1.351 | G22813T | S | 0.974 |
| B.1.351 | C25904T | ORF3a | 0.963 |
| B.1.351 | C25904T | ORF3b | 0.963 |
| B.1.351 | C26456T | E | 0.993 |

### References


1. Tyson JR, James P, Stoddart D, Sparks N, Wickenhagen A, Hall G, Choi JH, Lapointe H, Kamelian K, Smith AD, Prystajecky N, Goodfellow I, Wilson SJ, Harrigan R, Snutch TP, Loman NJ, Quick J. 2020. Improvements to the ARTIC multiplex PCR method for SARS-CoV-2 genome sequencing using nanopore. bioRxiv 2020.09.04.283077.
2. Freed NE, Vlková M, Faisal MB, Silander OK. 2020. Rapid and inexpensive whole-genome sequencing of SARS-CoV-2 using 1200 bp tiled amplicons and Oxford Nanopore Rapid Barcoding. Biol Methods Protoc 5.
3. Bushnell B. 2014. BBMap: a fast, accurate, splice-aware aligner. Lawrence Berkeley National Lab.(LBNL), Berkeley, CA (United States). Available at: https://sourceforge.net/projects/bbmap/
4. Joshi NA, Fass J. 2011. Sickle: A sliding-window, adaptive, quality-based trimming tool for FastQ files (Version 1.33)[Software]. Available at https://github.com/najoshi/sickle.
5. Li H. 2018. Minimap2: pairwise alignment for nucleotide sequences. Bioinformatics 34:3094–3100.
6. Li H, Handsaker B, Wysoker A, Fennell T, Ruan J, Homer N, Marth G, Abecasis G, Durbin R. 2009. The Sequence Alignment/Map format and SAMtools. Bioinformatics 25:2078–2079.
7. Masella AP, Lalansingh CM, Sivasundaram P, Fraser M, Bristow RG, Boutros PC. 2016. BAMQL: a query language for extracting reads from BAM files. BMC Bioinformatics 17:305.
8. BC Center for Disease Control. 2021. Weekly update on Variants of Concern (VOC). May 6, 2021.
9. D'Aoust PM, Mercier E, Montpetit D, Jia J-J, Alexandrov I, Neault N, Baig AT, Mayne J, Zhang X, Alain T, Langlois M-A, Servos MR, MacKenzie M, Figeys D, MacKenzie AE, Graber TE, Delatolla R. 2021. Quantitative analysis of SARS-CoV-2 RNA from wastewater solids in communities with low COVID-19 incidence and prevalence. Water Res 188:116560.
